# Pharmacological Management of Acute Spinal Cord Injury: A longitudinal multi-cohort observational study

**DOI:** 10.1101/2021.05.28.21257947

**Authors:** Catherine R. Jutzeler, Lucie Bourguignon, Bobo Tong, Elias Ronca, Eric Bailey, Noam Y. Harel, Fred Geisler, Adam R. Ferguson, Brian K. Kwon, Jacquelyn J. Cragg, Lukas Grassner, John L.K. Kramer

## Abstract

**Background:** Nearly every individual sustaining traumatic spinal cord injury receives multiple types and classes of medications to manage a litany of secondary complications. Prior clinical studies and evidence from animal models suggest that several of these medications could enhance or impede endogenous neurological recovery. However, there is a knowledge gap surrounding the spectrum of pharmacologic agents typically administered in the routine management of spinal cord injury.

**Objective:** To systematically determine the types of medications commonly administered, alone or in combination, in the acute to subacute phase of spinal cord injury.

**Methods:** We conducted an analysis of two largescale cohorts (the Sygen interventional trial and the SCIRehab observational cohort study) to determine what constitutes “ standards of acute pharmacological care” after spinal cord injury. Concomitant medication use, including dosage, timing and reason for administration, was tracked. Descriptive statistics were used to describe the medications administered within the first 60 days after spinal cord injury.

**Results:** Across 2040 individuals with spinal cord injury, 775 unique medications were administered within the two months after injury. On average, patients enrolled in the Sygen trial received 9.9 ± 4.9 (range 0-34), 14.3 ± 6.3 (range 1-40), 18.6 ± 8.2 (range 0-58), and 21.5 ± 9.7 (range 0-59) medications within the first 7, 14, 30, and 60 days post-injury, respectively. Patients enrolled in the SCIRehab cohort study received on average 1.7 ± 1.7 (range 0-11), 3.7 ± 3.7 (range 0-24), 8.5 ± 6.3 (range 0-42), and 13.5 ± 8.3 (range 0-52) medications within the first 7, 14, 30, and 60 days post-injury, respectively. Polypharmacy was commonplace (up to 43 medications per day per patient). Approximately 10% of medications were administered acutely as prophylaxis (e.g., against the development of pain or infections).

**Conclusions:** To our knowledge, this was the first time acute pharmacological practices have been comprehensively examined after spinal cord injury. Our study revealed a high degree of polypharmacy in the acute stages of spinal cord injury, with potential to both positively and negatively impact neurological recovery. This data may provide key insight to achieve better understanding of how the acute pharmacological management of spinal cord injury affects long-term recovery. All results can be interactively explored on the *R*_*X*_*SCI* web site (https://jutzelec.shinyapps.io/RxSCI/) and GitHub repository (https://github.com/jutzca/Acute-Pharmacological-Treatment-in-SCI/).

## INTRODUCTION

Traumatic spinal cord injury is a neurological condition associated with varying degrees of paralysis, sensory impairments, and autonomic deficits.^1^ At present, there are no pharmacological interventions available to enhance the extent a person neurologically or functionally recovers from acute spinal cord injury.^2,3^ In the absence of such an intervention, acute care chiefly focuses on managing the neurological sequela of spinal cord injury as well as emerging secondary complications, including infections, pain, and cardiovascular problems.^4^

Recent observational studies have reported a potential beneficial effect of acutely administered gabapentionoid medications (but not other anticonvulsants) on long-term neurological outcomes after spinal cord injury.^5–7^ Subsequent preclinical studies demonstrated a potential gabapentionoids-meditated mechanism for enhanced recovery, as well as confirmed behavioral benefits in animal models.^8,9^ While efficacy awaits confirmation in prospective clinical trials, these collective observations point to the promise of a reverse translational approach (bedside-to-bench) to restore neurological function after spinal cord injury. Identifying other opportunities for drug repurposing depends, in part, on knowledge regarding specific medications commonly administered in the acute phase. Additionally, if promising pharmacologic agents are to be proposed for human evaluation in clinical trials of acute spinal cord injury, it is important to consider the spectrum of other concomitant medications that are routinely administered in the care of these patients, as they may have known interactions with the promising agent in question. The aim of this study was to comprehensively characterize the pharmacological management of acute spinal cord injury. To this end, we leveraged available clinical trial and observational study data to determine the type, timing, and reason for administration associated with common medications.

## METHODS

### Study design

The design and reporting of this analysis adhered to the STROBE guidelines for observational studies.^10^

### Data source and cohort definition

To quantify medications commonly administered in the acute management of spinal cord injury, we analyzed two sources of data. Both sources represent collections of data from the United States, the first (i.e., trial) between 1992 and 1998 and the second (i.e., observational) from 2007-2009.

The first source comprised details of concomitant medications administered in a clinical trial - the Sygen trial - delivering GM-1 ganglioside in acute spinal cord injury.^2,11^ The Sygen trial was a randomized, prospective, phase III, placebo controlled, multi-center study testing the efficacy of GM-1 ganglioside therapy in acute, traumatic spinal cord injury.^2,11^ Full design, recruitment, and enrollment details have been published previously.^13^ Briefly, to be included in the Sygen trial patients were required to have at least one lower extremity with a substantial motor deficit. Patients with spinal cord transection or penetration were excluded, as were patients with a cauda equina, brachial or lumbosacral plexus, or peripheral nerve injury. Multiple trauma cases were included as long as they were not so severe as to preclude neurologic evaluation. Patients with major head trauma, major chest trauma, or intubation were also excluded. With 797 enrolled patients followed over the first year following injury, the Sygen trial was the largest clinical trial ever conducted in the field of spinal cord injury. The Sygen trial, which followed patients over the first year following injury, was clinically active from 1992 to 1998, and showed no differences between treatment and placebo groups in terms of neurological recovery.^2^ The negative finding of the Sygen study is considered Class I Medical Evidence by the spinal cord injury Committee of the American Association of Neurological Surgeons (AANS) and the Congress of Neurological Surgeons (CNS).^12,13^ Subsequent analyses of the Sygen data have been performed to characterize the trajectory and extent of spontaneous recovery from acute spinal cord injury.^14,15^

Our second source of data was from a large, observational study (i.e., SCIRehab), which abstracted information pertaining to medication use in the acute phase of spinal cord injury from patient medical records.^16^ The SCIRehab study enrolled, upon consent, individuals aged ≥12 years with traumatic spinal cord injury who were rehabilitated at six participating rehabilitation centers from 2007 through 2009.^17^ Participating centers included Rocky Mountain Regional Spinal Injury System at Craig Hospital, Shepherd Center, Atlanta GA; Rehabilitation Institute of Chicago, Chicago, IL; Carolinas Rehabilitation, Charlotte, NC; the Mount Sinai Medical Center, New York, NY; and National Rehabilitation Hospital, Washington, DC. Patients were followed for the first-year post-injury and were excluded if they spent two or more weeks at a non-participating rehabilitation center. Details of more than 460,000 interventions provided to 1500 patients were documented by over 1000 clinicians at the six participating centers. Patient demographics and injury characteristics were extracted from the patient medical record (part of the National Institute on Disability and Rehabilitation Research Spinal Cord Injury Model Systems Form I). Design, recruitment, inclusion criteria, and enrollment details have been previously described in detail.^17^

To be included in our study, information on medications administered needed to be available for the patients.

### Commonly administered medications

In the Sygen trial, alongside serious adverse events, concomitant medication information was routinely tracked following standardized case report forms by trained examiners in clinical trials as a measure of safety. For each concomitant medication administered during the trial, the reason for administration, dosage, dosing (i.e., start and end date, frequency), and reason for conclusion were recorded. It was also documented in case medications were administered for prophylactic reasons (e.g., to prevent deep vein thrombosis). Note that, although patients were randomized to GM-1 ganglioside therapy, individuals were not randomized to any concomitant medication administered and were managed according to the conventional care protocols of the enrolling center. The SCIRehab study documented the use of all commonly administered medications. For each medication administered, route, dosage and dosing (i.e., start and end date, frequency) were abstracted directly from medical records. However, medication indication was not recorded.

### Medication data cleaning and organizing

Medication data from the Sygen trial and SCIRehab study were separately cleaned and organized. From the medication files, which exist for each patient in the Sygen trial and SCIRehab, we extracted generic medication name and information on dosing (i.e., start and end date, frequency). As information on medication indication (i.e., reasons for administering a medication) was not entered in a standardized fashion during data collection, we classified the medication indication according to the Common Terminology Criteria for Adverse Events (CTCAE).^18^ Briefly, each indication was assigned to a System Organ Class (SOC),^18^ the highest level of the MedDRA hierarchy.^19^ The SOC is identified by anatomical or physiological system, etiology, or purpose (e.g., SOC Investigations for laboratory test results) and comprises 26 different categories. We added a separate class for trauma-related pain (i.e., nociceptive and neuropathic). The rationale for this amendment stems from the fact that the CTCAE does not sufficiently cover this category.

### Assessment of blood brain barrier (BBB) permeability

Leveraging the information from the DrugBank database (www.drugbank.ca), we determined which medications have the ability to cross the blood brain barrier. In case corresponding information was missing in the DrugBank, we searched PubMed for evidence.

### Statistical analysis and data visualization

R Statistical Software version 3.6.3 (Running under: macOS Mojave 10.14.4) was used for all analyses and to visualize the results. Descriptive statistics (mean, standard deviation, ranges, and proportions) were used to describe the patients’ demographics, injury characteristics, and medication information. For the latter, this included the number and type of medications administered, reason for administration, and how many medications each patient received per day (i.e., point prevalence). Type and frequency of medications that were administered prophylactically were also computed.

### **Interactive Web Platform** *R*_*X*_*SCI*

In order to enable the spinal cord injury community, researchers, authorities, and policymakers to fully explore the data and results of this study (and beyond), we developed the freely available and open source *R*_*X*_*SCI* web platform. *R*_*X*_*SCI* was implemented with the *Shiny* framework,^20^ which combines the computational power of the free statistical software R^21^ with friendly and interactive web interfaces. Both, the front- and back-end of *R*_*X*_*SCI* have been built using the *shiny dashboard* package.^22^ *R*_*X*_*SCI* is available as an online application and is hosted at https://jutzelec.shinyapps.io/RxSCI/ and can be accessed via any web browser on any device (e.g., desktop computers, laptops, tablets, smartphones). *R*_*X*_*SCI* is published under the BSD 3-Clause License. The source code of *Neurosurveillance* is available through Github at https://github.com/jutzca/Acute-Pharmacological-Treatment-in-SCI/tree/master/shinyapp.

### Data sharing and code availability

Full anonymized data of both data sources will be shared at the request from any qualified investigator (please contact the *Corresponding Author*). The code for the data analysis and visualization is available in our GitHub repository (https://github.com/jutzca/Acute-Pharmacological-Treatment-in-SCI/).

### Standard Protocol Approvals, Registrations, and Patient Consents

Approval for this study (secondary analysis) was received by an institutional ethical standards committee on human experimentation at the University of British Columbia. The original Sygen clinical trial (results published elsewhere) also received ethical approval, but was conducted before clinical trials were required to be registered (i.e., no clinicaltrial.gov identifier available)^2,23^. Each participating center of the SCIRehab study received institutional review board approval for this study and obtained informed consent from each patient (or their parent/guardian).

## RESULTS

### Patient Characteristics and Summary Statistics

We included 797 and 1243 patients from the Sygen clinical trial and SCIRehab observational study, respectively. While all patients from the Sygen study were included in our analysis, we had to exclude 275 patients from the SCIRehab study due to missing data on medications (n=241) or absence spinal cord injury (i.e., AIS E, cauda equine or peripheral nervous system injuries, n=14). In both cohorts, the ratio between male and female patients was approximately 4:1, the majority of the patients were injured at the cervical levels (Sygen: 75.2%; SCIRehab: 60.4%) and sustained a motor complete injury (Sygen: 65.7%; SCIRehab: 65.6%). The most frequent cause of injury was car accidents (Sygen: 47.9%; SCIRehab: 35.5%) followed by falls (Sygen: 16.2%; SCIRehab: 24.1%). Detailed description of both cohorts is provided in **Table 1**.

**Table 1.**
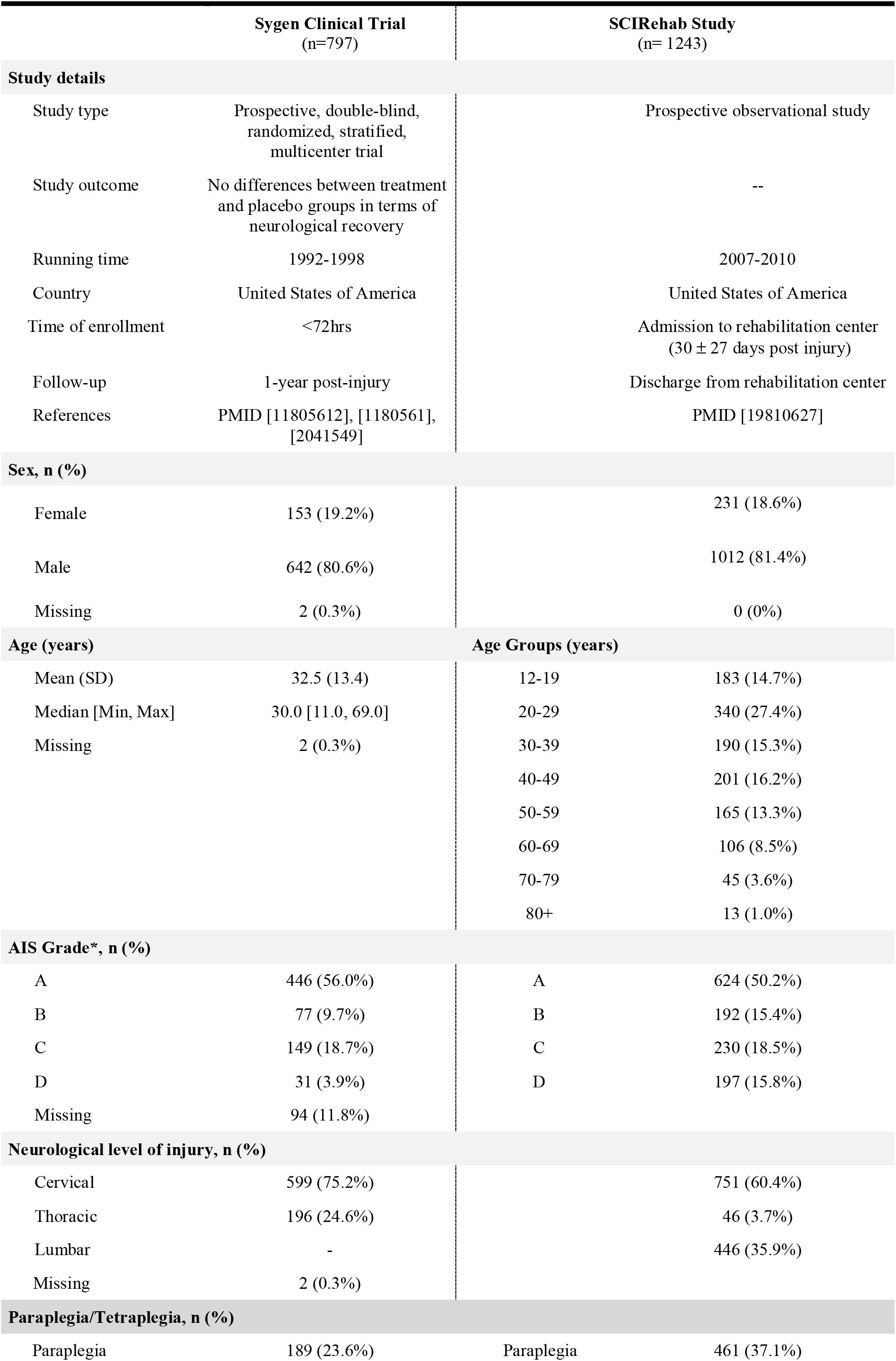
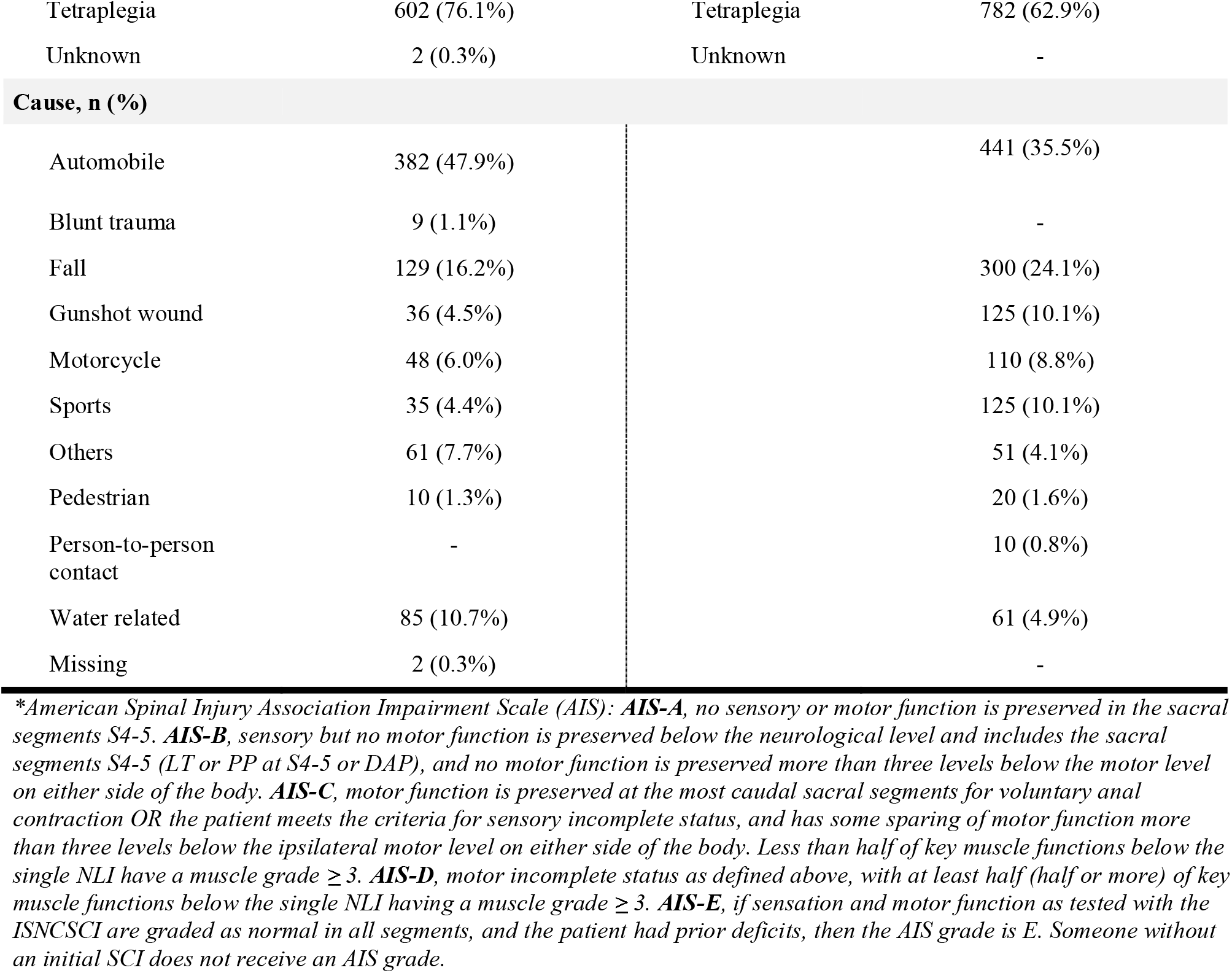
Demographics and injury characteristics of the included cohorts.

|  | **Sygen Clinical Trial**<br>(n=797) | **SCIRehab Study**<br>(n= 1243) |
| --- | --- | --- |
| **Study details** |  |  |
| Study type | Prospective, double-blind, randomized, stratified, multicenter trial | Prospective observational study |
| Study outcome | No differences between treatment and placebo groups in terms of neurological recovery | -- |
| Running time | 1992-1998 | 2007-2010 |
| Country | United States of America | United States of America |
| Time of enrollment | <72hrs | Admission to rehabilitation center (30 ± 27 days post injury) |
| Follow-up | 1-year post-injury | Discharge from rehabilitation center |
| References | PMID [11805612], [1180561], [2041549] | PMID [19810627] |
| **Sex, n (%)** |  |  |
| Female | 153 (19.2%) | 231 (18.6%) |
| Male | 642 (80.6%) | 1012 (81.4%) |
| Missing | 2 (0.3%) | 0 (0%) |
| **Age (years)** |  | **Age Groups (years)** |
| Mean (SD) | 32.5 (13.4) | 12-19 183 (14.7%) |
| Median [Min, Max] | 30.0 [11.0, 69.0] | 20-29 340 (27.4%) |
| Missing | 2 (0.3%) | 30-39 190 (15.3%) |
|  |  | 40-49 201 (16.2%) |
|  |  | 50-59 165 (13.3%) |
|  |  | 60-69 106 (8.5%) |
|  |  | 70-79 45 (3.6%) |
|  |  | 80+ 13 (1.0%) |
| **AIS Grade\*, n (%)** |  |  |
| A | 446 (56.0%) | A 624 (50.2%) |
| B | 77 (9.7%) | B 192 (15.4%) |
| C | 149 (18.7%) | C 230 (18.5%) |
| D | 31 (3.9%) | D 197 (15.8%) |
| Missing | 94 (11.8%) |  |
| **Neurological level of injury, n (%)** |  |  |
| Cervical | 599 (75.2%) | 751 (60.4%) |
| Thoracic | 196 (24.6%) | 46 (3.7%) |
| Lumbar | - | 446 (35.9%) |
| Missing | 2 (0.3%) |  |
| **Paraplegia/Tetraplegia, n (%)** |  |  |
| Paraplegia | 189 (23.6%) | Paraplegia 461 (37.1%) |

|  |  |  |  |
| --- | --- | --- | --- |
| Tetraplegia | 602 (76.1%) | Tetraplegia | 782 (62.9%) |
| Unknown | 2 (0.3%) | Unknown | - |
| **Cause, n (%)** |  |  |  |
| Automobile | 382 (47.9%) |  | 441 (35.5%) |
| Blunt trauma | 9 (1.1%) |  | - |
| Fall | 129 (16.2%) |  | 300 (24.1%) |
| Gunshot wound | 36 (4.5%) |  | 125 (10.1%) |
| Motorcycle | 48 (6.0%) |  | 110 (8.8%) |
| Sports | 35 (4.4%) |  | 125 (10.1%) |
| Others | 61 (7.7%) |  | 51 (4.1%) |
| Pedestrian | 10 (1.3%) |  | 20 (1.6%) |
| Person-to-person contact | - |  | 10 (0.8%) |
| Water related | 85 (10.7%) |  | 61 (4.9%) |
| Missing | 2 (0.3%) |  | - |

*\*American Spinal Injury Association Impairment Scale (AIS): **AIS-A**, no sensory or motor function is preserved in the sacral segments S4-5. **AIS-B**, sensory but no motor function is preserved below the neurological level and includes the sacral segments S4-5 (LT or PP at S4-5 or DAP), and no motor function is preserved more than three levels below the motor level on either side of the body. **AIS-C**, motor function is preserved at the most caudal sacral segments for voluntary anal contraction OR the patient meets the criteria for sensory incomplete status, and has some sparing of motor function more than three levels below the ipsilateral motor level on either side of the body. Less than half of key muscle functions below the single NLI have a muscle grade $\geq 3$ . **AIS-D**, motor incomplete status as defined above, with at least half (half or more) of key muscle functions below the single NLI having a muscle grade $\geq 3$ . **AIS-E**, if sensation and motor function as tested with the ISNCSCI are graded as normal in all segments, and the patient had prior deficits, then the AIS grade is E. Someone without an initial SCI does not receive an AIS grade.*

### Acute pharmacological management after spinal cord injury

In total, 489 (trial) and 575 (observational study) unique medications were administered over the course of 60 days after spinal cord injury. More than a third (n=289 [∼37.3%]) of the medications administered were common to both data sources (for details see **Table e1**). Medications were administered to manage secondary complications arising from 20 different system organ classes or to facilitate surgical and medical procedures (**Figure 1A**). On average, patients enrolled in the Sygen trial received 9.9 ± 4.9 (range 0-34), 14.3 ± 6.3 (range 1-40), 18.6 ± 8.2 (range 0-58), and 21.5 ± 9.7 (range 0-59) medications within the first 7, 14, 30, and 60 days post-injury, respectively (**Figure 1B**). Patients enrolled in the SCIRehab cohort study received on average 1.7 ± 1.7 (range 0-11), 3.7 ± 3.7 (range 0-24), 8.5 ± 6.3 (range 0-42), and 13.5 ± 8.3 (range 0-52) medications within the first 7, 14, 30, and 60 days post-injury, respectively (**Figure 1C**). The disparity between Sygen and SCIRehab in the first month post injury can be attributed to different time-points of patient enrollment, with the Sygen trial enrolling patients within 72 hours, compared to SCIRehab, which enrolled patients within days or weeks of injury (**Table 1**). As a result, medications for first-line trauma management (e.g., nitroglycerin, dopamine) as well as surgical and medical procedures (e.g., isoflurane, vecuronium bromide) are only captured by the Sygen trial. Acetaminophen (analgesic, n = 674 patients), morphine (analgesic, n = 664 patients), and heparin (anticoagulant, n = 505 patients) were the three most commonly administered medications in the Sygen trial (**Figure 1D**). Similarly, in the SCIRehab study, the analgesic acetaminophen (n = 924 patients) was the most commonly administered medication, followed by the laxative docusate (n = 862 patients) and the analgesic combination medicine acetaminophen & oxycodone (n = 603 patients) (**Figure 1E**).

**Figure 1.**
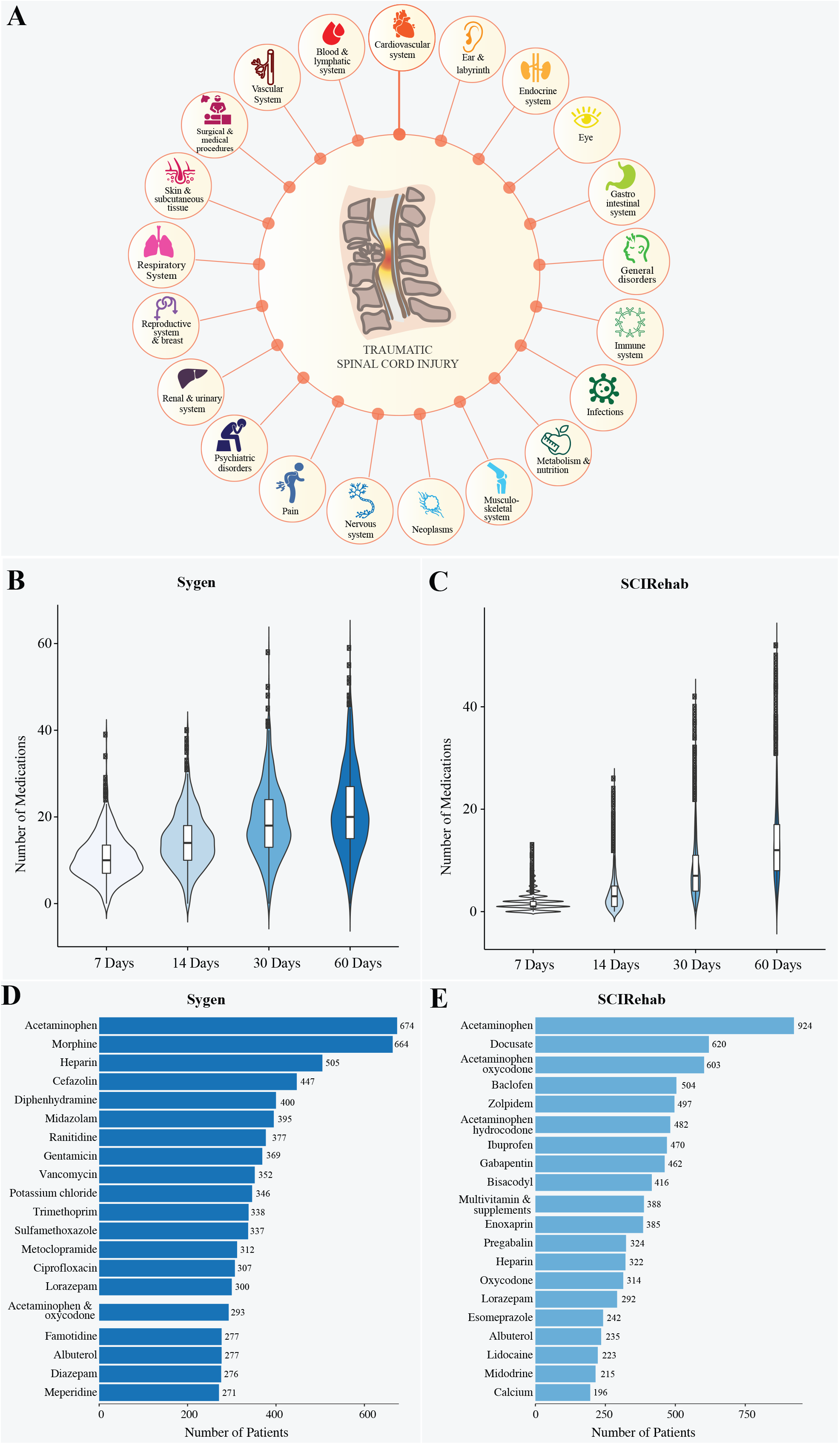
Pharmacological management of acute spinal cord injury. **(A) Secondary complications.** Spinal cord injury is associated with a large number of secondary complications that arise from 20 organ systems as defined by Common Terminology Criteria for Adverse Events (CTCAE) published by U.S. Department of Health and Human Services.^26^ Many medications were also administered to facilitate medical and surgical procedures, such as decompression surgeries, laminectomy, and computer tomography scans. **(B) Number of medications administered** to patients enrolled in the Sygen trial within the first 7, 14, 30, and 60 days post-injury. **(C) Number of medications administered** to patients enrolled in the SCIRehab study within the first 7, 14, 30, and 60 days post-injury. **(D) Frequency of medications administered**. The majority of patients enrolled in the Sygen trial received acetaminophen, morphine, and heparin to treat secondary complications, such as pain and deep venous thrombosis. **(E) Frequency of medications administered**. Pain killers (acetaminophen and Acetaminophen oxycodone) as well as the laxative docusate were among the most frequently administered medications in the SCIRehab study.

The majority of patients enrolled in the Sygen trial required medications to treat secondary complications arising from the gastrointestinal system (n = 752, 95.1%), pain (n = 742, 93.8%), infections (n = 737, 93.2%), and psychiatric issues (n = 650, 82.2%) (**Figure 2A, Table e2**). A total of 150, 99, and 93 unique medications were administered to treat a variety of secondary complications arising from infections, respiratory system, and gastrointestinal system, respectively. Moreover, pain (e.g., musculoskeletal), gastrointestinal complications (e.g., heartburn, ulcers), and infections (i.e., bacteria, viral, and fungal) were the most frequently managed problems (**Figure 2B, Table e3**). This was also true when stratifying for injury severity (AIS grades, **Table e4**). While infections were mainly treated with antibiotics, antifungal, and antiviral medications depending on their nature, complications arising from gastrointestinal tract were targeted with analgesics, antibiotics, antacids, antiulcer, anti-anemics, anticholinergics, anticonvulsants, and antispasmodics (see detailed overview in **Table e5**).

**Figure 2.**
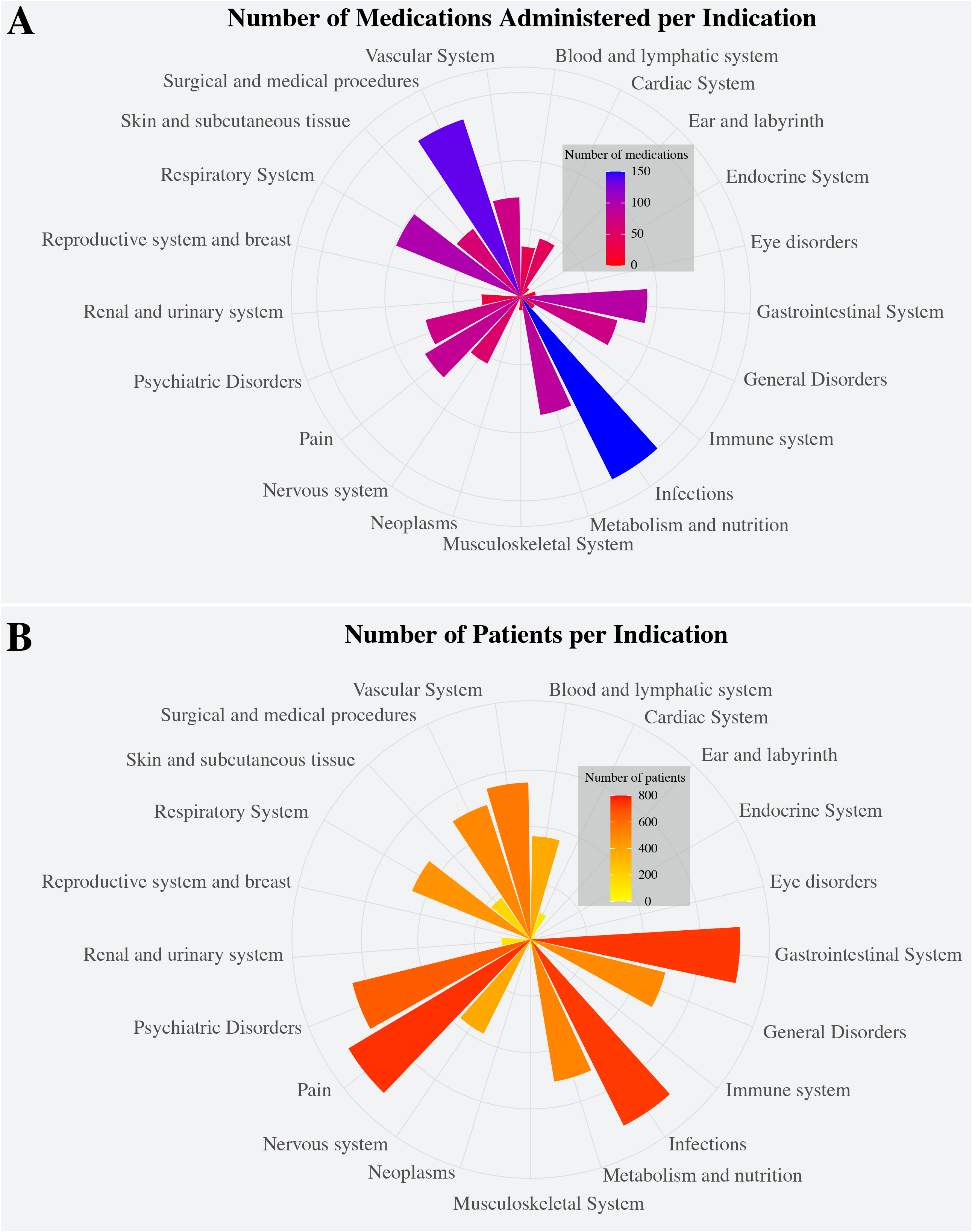
Indication of medications administered. **A) Number of unique medications administered per organ system for patients enrolled in the Sygen clinical trial.** Note the diversity of medications administered within each category of complications. For instance, over 100 different medications were administered to treat infections and infestations as well as for surgical and medical procedures. **(B) Number of patients of the Sygen clinical trial that required treatment per organ system**. The three most frequently treated secondary complications were pain, gastro-intestinal system disorders, as well as infections. The SCIRehab database did not track the indications for which medications were prescribed.

### Polypharmacy

As illustrated in **Figures 3**, polypharmacy was commonplace. Almost every patient enrolled the Sygen trial or the SCIRehab study received multiple medications per day (**Figure 3A**). Patients with more severe injuries (AIS A and B) received slightly more medications per day than those with less severe injuries (AIS D). The number of medications administered per day per patient ranged between 1 and 30 for patients enrolled in Sygen trial (**Figure 3B**) and between 1 and 43 for patients enrolled in the SCIRehab study (**Figure 3B**). Individual patient examples of the extend of polypharmacy is shown in **Figure 3C**. The complexity of the combination of medications administered is illustrated in **Figure 3D**. In the Sygen trial, the three most common combinations of medications were acetaminophen and morphine (n = 164 patients), morphine and ranitidine (n = 128 patients), as well as acetaminophen and heparin (n =123 patients). In the SCIRehab study, acetaminophen and acetaminophen oxycodone was the most common combination of medications (n = 480 patients), followed by acetaminophen and acetaminophen hydrocodone (n = 407 patients) as well as acetaminophen and ibuprofen (n = 346 patients.) The complexity of the combination of medications administered to patients in the SCIRehab study is illustrated in **Figure 3E**.

**Figure 3.**
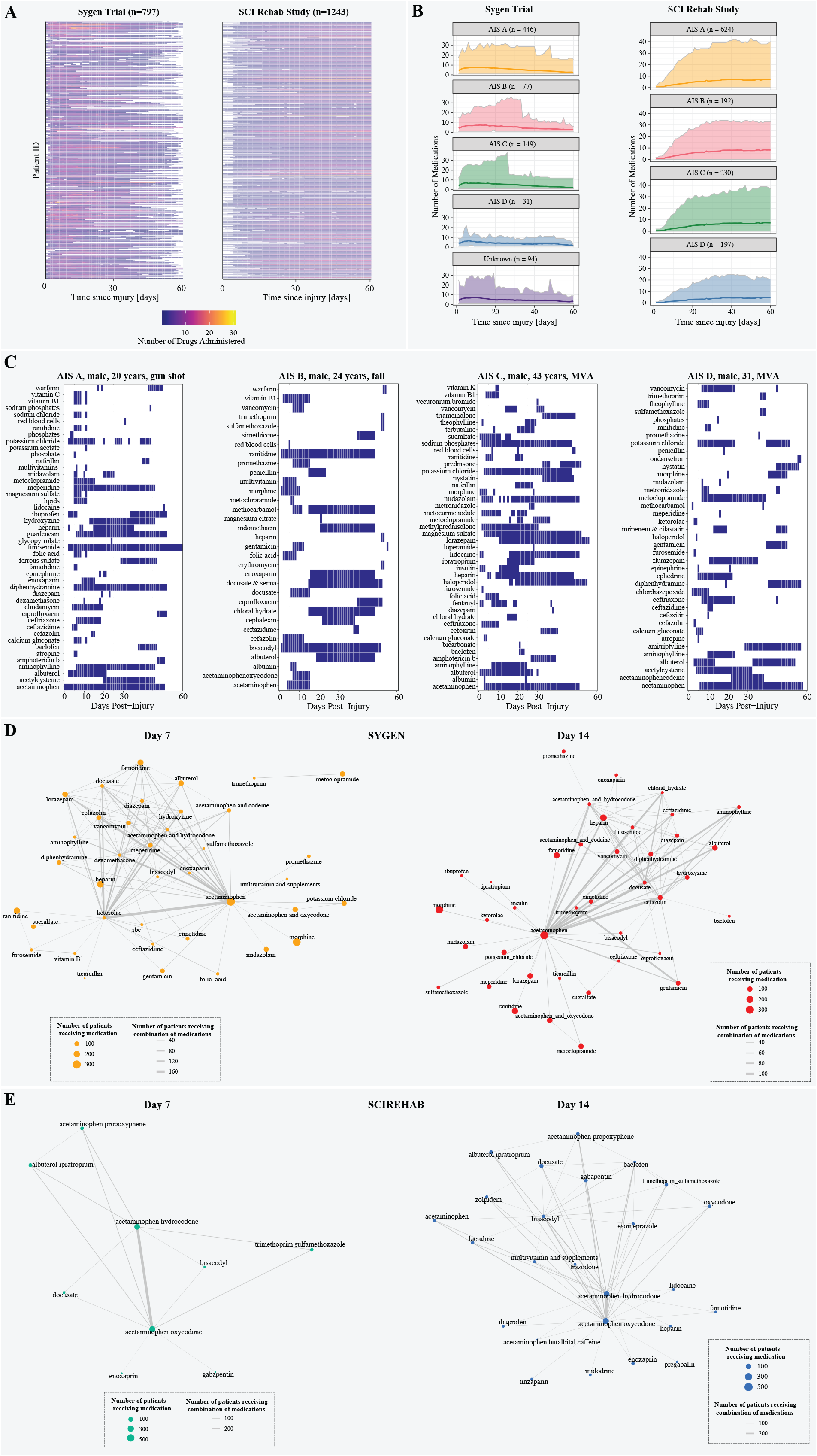
Polypharmacy. **(A) Point prevalence of commonly administered medications.** The number of medications administered per day per patient in the first 60 days post injury varied between 1 and 30 for the clinical trial and between 1 and 43 in the observational study. Each line represents one patient and the color white indicates that no medication was administered or no data was available for that time period. **(B) Daily average number of medications administered**. Patients with motor complete injuries (AIS A and B) received on average more medications per day compared to patients with motor incomplete injuries. The range medications administered varies quite drastically. The dashed line denotes the average number of medications and the solid lines the minimum and maximum number of medications, respectively. Patients with no information on AIS grades at baseline were grouped together in the category ‘unknown’. **(C) Examples longitudinal medication profiles for four patients in the first 60 days post injury**. Polypharmacy was commonplace across different injury severities and aetiologies. The pattern of medication administration varied between continuous, intermittent, and single-use indications. Medications were often co-administered bearing a high risk of pharmacological interactions between medications. While some are well-understood, the majority of these interactions (particularly combinations of three and more medications) have not yet been explored. **(D) Network of medications administered in combination to patients enrolled in the Sygen trial**. The nodes of the network represent the medications. The size of the nodes represents the number of patients that have received this particular medication on day 7 or 14, respectively. Medications that were administered together on a specific day, either 7 or 14, are connected via an edge. The width of the edge represents the number of patients that have received the two medications (acetaminophen and ketorolac) in combination on the day of interest. **(E) Network of medications administered in combination to patients enrolled in the SCIRehab study**. The nodes of the network represent the medications. The size of the nodes represents the number of patients that have received this particular medication on day 7 or 14, respectively.

### Blood brain barrier (BBB) permeability

Out of the 775 unique medications, 59.4% (n = 460) have the ability to cross the BBB while 20.6% (n = 160) are not permeable for the BBB. No information regarding the BBB permeability could be found for the remaining 20.0% (n = 155). Detailed information on the permeability can be found in **Table e6**. Drugs that cross the BBB may be more likely to have effects (positive or negative) on neural recovery pathways after injury.

### Prophylactic administration of medications

Approximately 10% (n = 2,838) of all recorded indications in the Sygen trial (**Figure 4A**) were labelled ‘prophylactic’ or ‘preventative’. A total of 137 unique medications were administered for prophylactic treatment to prevent a wide range of secondary complications (**Figure 4B**). The major medication groups included antihistamines (ranitidine, famotidine), anticoagulants (heparin, warfarin), and antibiotics (cefazolin, gentamicin) for the prevention of secondary complications arising from the gastrointestinal system (e.g., heart burn, gastric ulcers), blood and vasculature system (e.g., deep vein thrombosis), and infections, respectively (**Figure 4C**). The majority of patient enrolled in the Sygen trial (n = 666 [83.6%]) received prophylactic treatments (mean_medications/patient_ = 3 [range 1-21]; mean_indications/patient_ = 4.3 [range 1-33]) (**Figure 4D**). **Table e7** provides a comprehensive overview of all medications (and their respective indications) that were used for prophylactic treatment.

**Figure 4.**
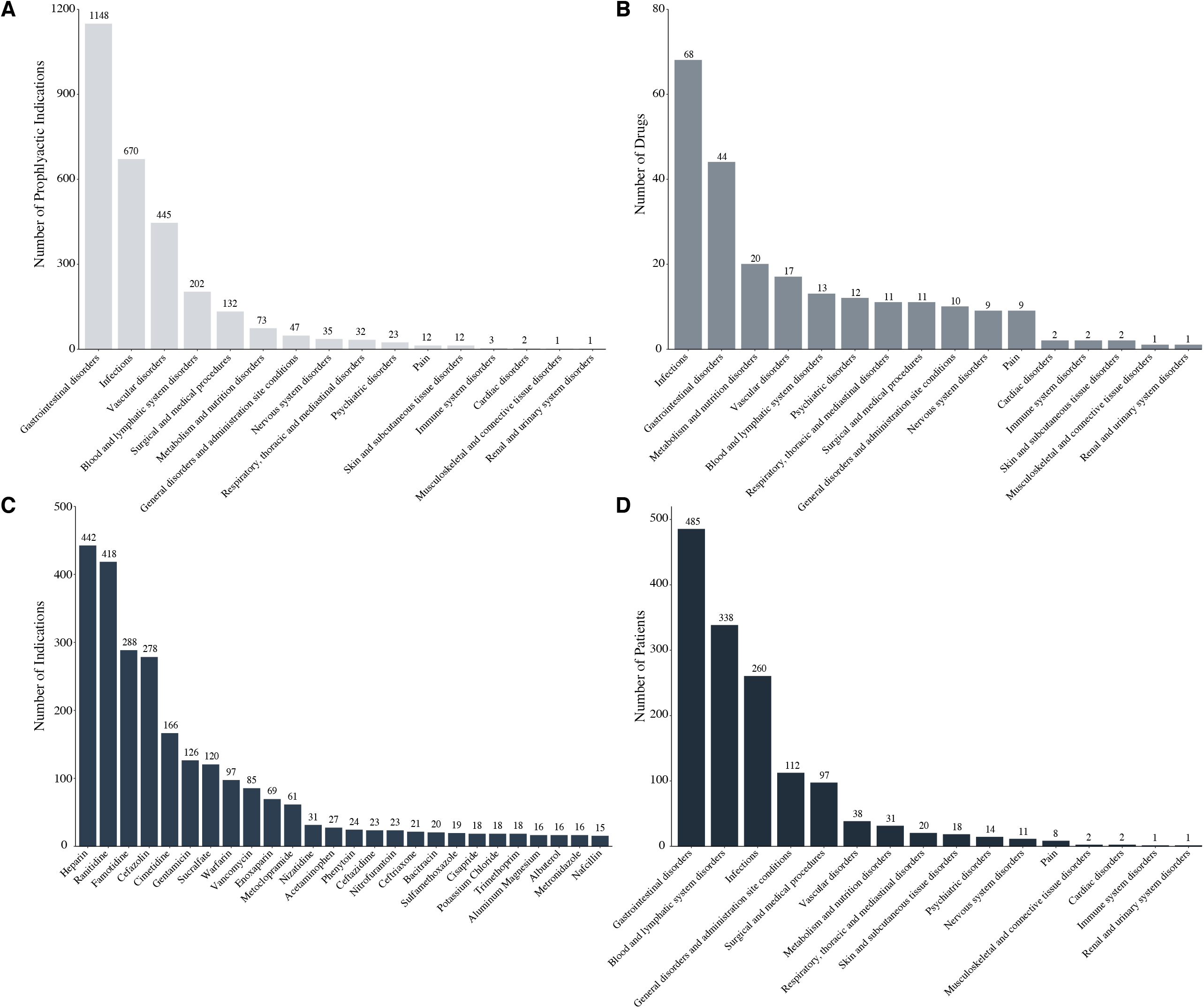
Prophylactic pharmacological treatment to prevent secondary complications from occurring. **(A) Number of indications per organ system.** The majority of prophylactic indications were related to the gastrointestinal and vascular system as well as infections of all sorts. **(B) Number of unique medications administered to for disease prophylaxis. (C) Number of indications per medications**. Anticoagulants, antihistamines, and antibiotics were amongst the most frequently administered medication classes. **(D) Number of patients that received prophylactic treatment per organ system**. The majority of the patients enrolled in the Sygen trial (n=666 [83.6%]) received at least one medication for disease prophylaxis. The average number of medications per patient was 3 (range: 1-21) and average number of indications per patient was 4.3 (1-33).

### **Interactive Web Platform** *R*_*X*_*SCI*

The *Neurosurveillance* web platform is hosted online (https://jutzelec.shinyapps.io/RxSCI/) and contains three main data visualization parts: (1) epidemiological features, including demographics and injury characteristic; (2) information on the pharmacological treatment of spinal cord injury patients on daily basis, including medication administration patterns; and (3) visualization of the polypharmacy. All data from the Sygen clinical trial and the SCIRehab study, which was used in this study, can be explored in a customized fashion (e.g., customized selection of patient groups). The platform is configured such that existing or newly generated data sets can be added if they comply with GDPR

## DISCUSSION

The aim of the current study was to comprehensively evaluate pharmacological management practices in acute spinal cord injury. To this end, two large data sources were examined, one from a clinical trial and the other from an observational study. Our analysis revealed an incredibly high rate of polypharmacy spread over the course of the first 60 days’ post injury, which was administered to manage various health conditions arising directly or indirectly from acute spinal cord injury. Various medications were administered, including those that readily cross the BBB as well as the spinal cord blood barrier (e.g., pregabalin,^24^ morphine^25^) to manage the sequela of spinal cord injury (e.g., neuropathic pain), as well as other complex medical complications.

To our knowledge, this was the first time acute pharmacological practices have been comprehensively examined after spinal cord injury. Even considering its extreme and traumatic nature, the sheer number of medications administered in a short window of time after spinal cord injury, over the course of the two months, was remarkably high. This led to a very high degree of polypharmacy. For comparison, polypharmacy in other complex health conditions is generally considered more than five medications^26,27^ – the average for acute spinal cord injury patients was approximately double that threshold. While perhaps startling, there is no doubt that aggressive pharmacologic management is typically required due to the complexity of managing spinal cord injury requires aggressive pharmacological management. Nevertheless, the lack of attention paid to the question of “ neurological safety” (i.e., whether use of a medication or its interaction with other medications in the acute phase of injury will have long-term and detrimental neurological consequences) is surprising, as is the fact that few attempts have been made to discern potential beneficial (or detrimental) effects of medications that readily cross the BBB. Furthermore, one must consider potential interactions between the high number of clinically used concomitant medications with novel medications and biologics being trialed for improving recovery from spinal cord injury.

The limited knowledge about the potential effects of acutely administered medications on recovery in humans becomes all the more curious considering that a number of these medications alter outcomes in animal studies. As an example, pregabalin, a potent calcium channel blocker and anticonvulsant administered for neuropathic pain, has been repeatedly shown to benefit recovery after spinal cord injury in animal and human spinal cord injury.^5,6,8,9^ Detrimental effects were also observed for some medications, including opioids, which attenuated the recovery of locomotor function and exacerbated pathophysiological processes in rodent models of spinal cord injury.^28,29^ A detrimental opioid effect is in line with beneficial effects of naloxone (i.e., opioid antagonist),^30–32^ and is highly concerning in light of the fact that opioids are ubiquitously administered for pain management in the early stages of injury (to > 80% of the patients). While completely removing or restricting opioids would be highly problematic and present with serious ethical concerns (i.e., weighing the management of acute pain with long-term neurological effects), opioids were among medications commonly administered to prevent the onset of pain. This suggests that opioids, at least in a proportion of patients, were prescribed with the intention to prevent the onset of pain, despite a lack of evidence.^33^ Among these individuals, neurological recovery could perhaps be facilitated by minimizing the administration of opioids. Many other common medications (up to 10%) are prophylactically administered, including acetaminophen, cefazolin, and famotidine for pain/fever, infection, and ulcer prophylaxis, respectively.

Despite years of use in clinical routine, safety information with respect to neurological outcomes of many concomitant medications is currently not available. This is highly concerning because fundamental assumptions of pharmacokinetics and -dynamics may not apply as in other (healthy) individuals.^34^ Alterations in physiology lead to prolonged absorption as a consequence of slowed gastric emptying and gastrointestinal motility,^35,36^ altered distribution due to leaky blood spinal cord barrier,^37–39^ hampered metabolism,^37,40,41^ and slowed excretion are hallmarks of this altered physiology.^39,41,42^ Examples of medications with changed pharmacokinetics are amikacin, baclofen, carbamazepine, cefotiam, ciprofloxacin, diazepam, diclofenac, doxycycline, ketamine, lorazepam, naproxen, and vancomycin. A major issue with these injury-induced modifications in pharmacokinetics is that some medications do not reach desired therapeutic effects, whereas others may reach potentially toxic levels. In addition to potential toxicity, also common side effects of medications (e.g., gastric emptying and gastrointestinal motility caused by opioids) may worsen the natural pathophysiology of injury. Post-marketing surveillance and risk assessment programs aim at detecting previously unrecognized positive or negative effects that may be associated with a medication - within *real-world* populations. To our knowledge, few of these studies have examined effects after spinal cord injury. An exemption is a recent study that established neurological safety profile of baclofen, an antispasmodic to treat debilitating muscle spasms.^43^ Cragg et al performed a secondary analysis of clinical trial data to provide data reaffirming that baclofen is neurologically, hepatically, and renally safe to use in patients sustaining a spinal cord injury.^43^ Complementing the existing safety profile, neurological safety medication profiles in the context of concomitant medications in real-world settings will enable health care providers to provide an informed, evidence-based response regarding the use of medications such as baclofen in the acute phase of spinal cord injury.

### Conclusion and implications for other neurological disorders

Our study revealed that in the management of acute spinal cord injury, there is a dramatic degree of polypharmacy that could potentially impact recovery and the potency of novel treatments of spinal cord injury. It should be noted that in the testing of novel drug agents in preclinical models of spinal cord injury, the experiments are typically designed to minimize (and of course standardize) the concomitant medications administered to the animals. Our analysis reveals how starkly different these experimental conditions are from clinical reality. Spinal cord injury is a complex condition and as such, the pharmacologic needs are understandably high. While we are not arguing for an arbitrary “ reduction” in the use of various medications in the management of these individuals, evaluating current standards of acute care and understanding what pharmacologic agents patients are typically exposed to does represent an intriguing alternative strategy to improve the lives of individuals with spinal cord injury, and at the least highlights the need for awareness when designing drug trials for the acute injury setting. Knowledge gained from our study has major implications for other diseases hallmarked by polypharmacy, including Parkinson’s disease,^44^ Alzheimer’s disease,^45^ Multiple Sclerosis,^46^ traumatic brain injury,^47,48^ cancer,^49^ and sepsis.^50^ Similar to spinal cord injury, these diseases are complex conditions associated with a wide range of symptoms (e.g., functional impairment) and secondary complications (e.g., gastrointestinal and cardiovascular complications, pain) necessitating pharmacological treatment – at times simultaneously. Many of these diseases are not yet curable, but effective disease modifying treatments that relieve symptoms, slow down disease progression, and improve quality of life are available.^51–54^ A cursory glance at the literature corroborates that the knowledge gap regarding the effect of commonly used medications on disease progression and their potential to alter the effectiveness of disease modifying treatments is not unique to spinal cord injury.

## Data Availability

Full anonymized data of both data sources will be shared at the request from any qualified investigator (please contact the Corresponding Author). The code for the data analysis and visualization is available in our GitHub repository (https://github.com/jutzca/Acute-Pharmacological-Treatment-in-SCI/).

## Acknowledgments

The authors would like to acknowledge the participating centers in the Sygen trial and SCIRehab network that were involved in the patient care and collection of data necessary for this study.

## Appendix 1: Authors

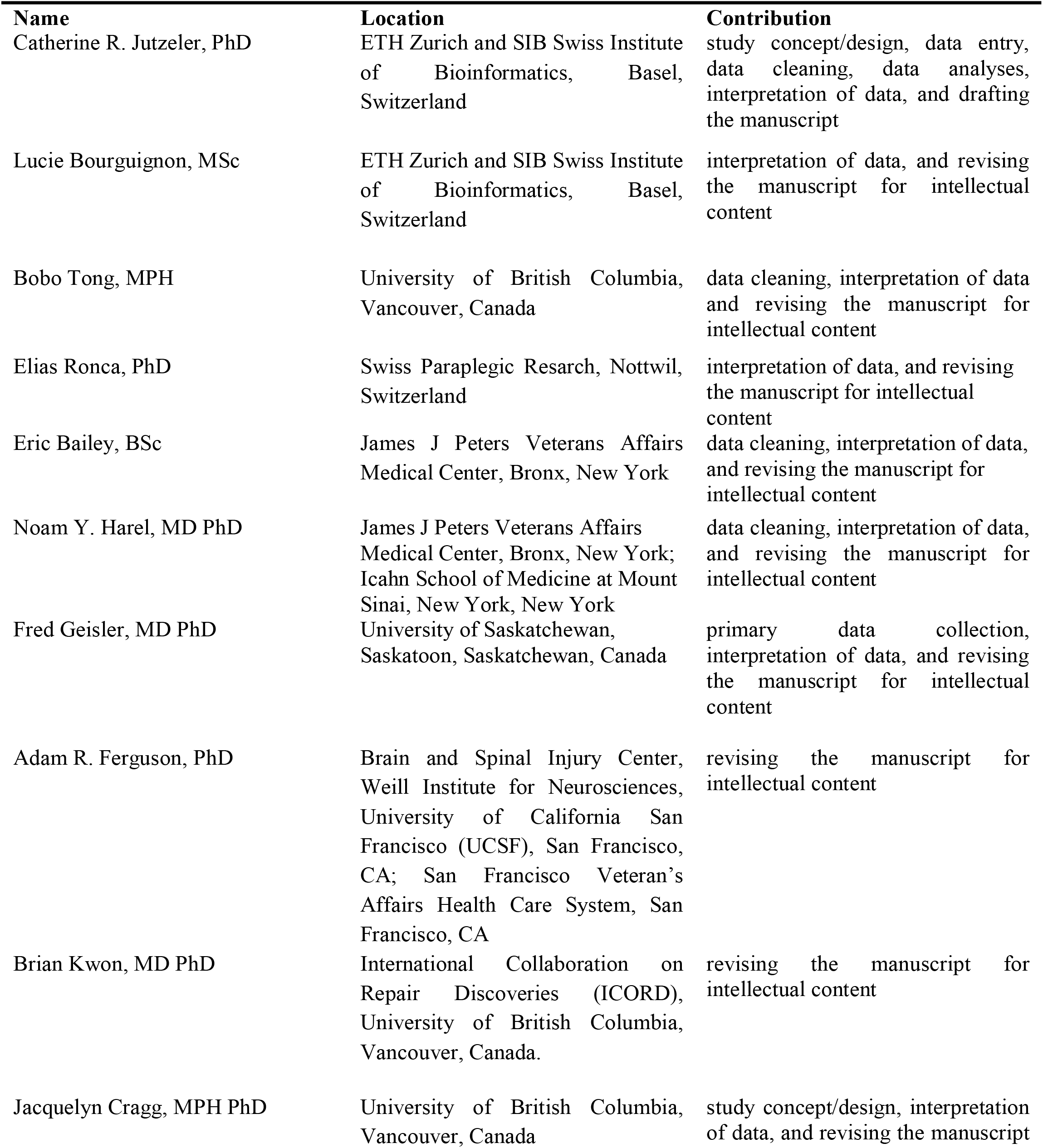
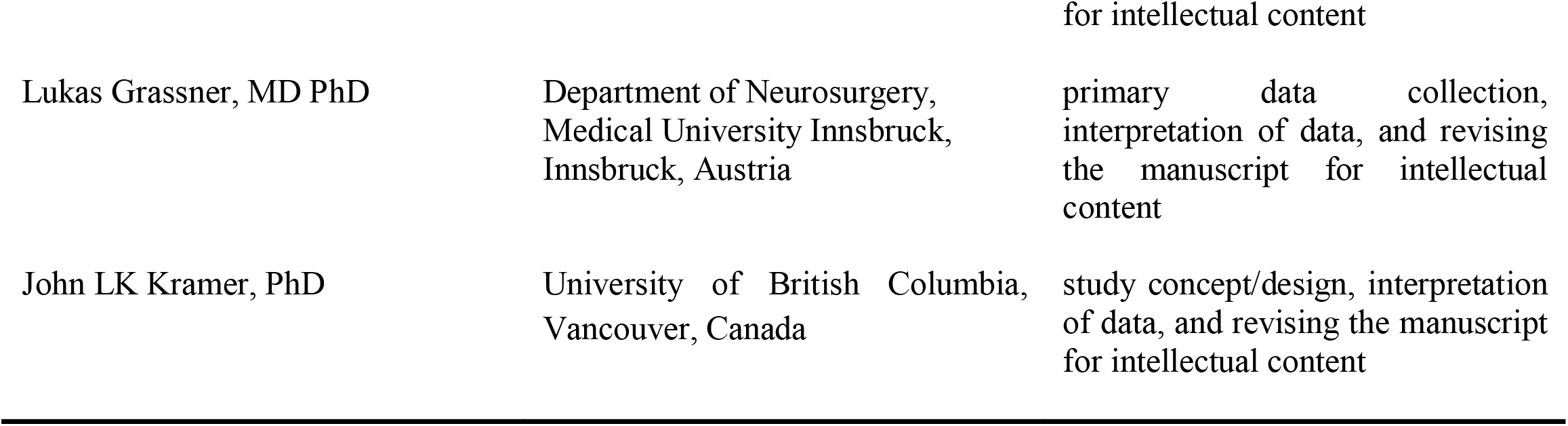

