## Supplementary material for "Pharmacological Management of Acute Spinal Cord Injury: A longitudinal multi-cohort observational study": Table e2

| **Supplementary Table 2.** Number of patients that required treatment for secondary complications arising from any of the 21 different organ systems or as preparation for surgical and medical procedures. | |
| --- | --- |
| **Indication** | **Number of patients receiving treatment** |
| Pain | 752 |
| Gastrointestinal System | 742 |
| Infections and infestations | 737 |
| Psychiatric Disorders | 650 |
| Vascular System | 556 |
| Metabolism and nutrition | 510 |
| Surgical and medical procedures | 501 |
| General Disorders | 491 |
| Respiratory System | 454 |
| Nervous system | 373 |
| Blood and lymphatic system | 366 |
| Skin and subcutaneous tissue | 181 |
| Renal and urinary system | 104 |
| Cardiac System | 100 |
| Immune system | 32 |
| Musculoskeletal System | 28 |
| Ear and labyrinth | 21 |
| Eye | 20 |
| Reproductive system and breast | 4 |
| Endocrine System | 3 |
| Neoplasms | 1 |
