## Supplementary material for "Pharmacological Management of Acute Spinal Cord Injury: A longitudinal multi-cohort observational study": Table e3

| **Supplementary Table 3.** Number of unique medications that were administered to treat secondary complications arising from 21 organ systems or for surgical and medical procedures. | |
| --- | --- |
| **Indication** | **Number of unique medications** |
| Infections and infestations | 150 |
| Surgical and medical procedures | 137 |
| Respiratory System | 99 |
| Gastrointestinal System | 93 |
| Metabolism and nutrition | 88 |
| Pain | 82 |
| General Disorders | 73 |
| Vascular System | 73 |
| Psychiatric Disorders | 72 |
| Skin and subcutaneous tissue | 61 |
| Nervous system | 55 |
| Cardiac System | 45 |
| Blood and lymphatic system | 37 |
| Renal and urinary system | 29 |
| Immune system | 12 |
| Eye disorders | 11 |
| Musculoskeletal System | 10 |
| Ear and labyrinth | 8 |
| Reproductive system and breast | 3 |
| Endocrine System | 2 |
| Neoplasms | 2 |
