## Supplementary material for "Pharmacological Management of Acute Spinal Cord Injury: A longitudinal multi-cohort observational study": Table e4

| **Supplementary Table 3.** Number of unique medications that were administered to treat secondary complications arising from 21 organ systems or for surgical and medical procedures. The stratification was performed according to the injury severity measured according to the AIS grades. | | |
| --- | --- | --- |
| **Baseline AIS Grade** | **Indication** | **n** |
| AIS-A | Infections and infestations | 136 |
|  | Surgical and medical procedures | 107 |
|  | Respiratory System | 86 |
|  | Gastrointestinal System | 76 |
|  | Metabolism and nutrition | 76 |
|  | General Disorders | 66 |
|  | Pain | 63 |
|  | Psychiatric Disorders | 58 |
|  | Vascular System | 55 |
|  | Nervous system | 43 |
|  | Skin and subcutaneous tissue | 41 |
|  | Blood and lymphatic system | 26 |
|  | Cardiac System | 26 |
|  | Renal and urinary system | 21 |
|  | Immune system | 8 |
|  | Eye | 6 |
|  | Ear and labyrinth | 5 |
|  | Musculoskeletal System | 5 |
|  | Endocrine System | 2 |
|  | Reproductive system and breast | 2 |
| AIS-B | Infections and infestations | 64 |
|  | Surgical and medical procedures | 46 |
|  | Pain | 42 |
|  | Gastrointestinal System | 38 |
|  | Psychiatric Disorders | 33 |
|  | Respiratory System | 30 |
|  | Metabolism and nutrition | 29 |
|  | Vascular System | 26 |
|  | General Disorders | 20 |
|  | Nervous system | 19 |
|  | Skin and subcutaneous tissue | 19 |
|  | Blood and lymphatic system | 11 |
|  | Cardiac System | 8 |
|  | Renal and urinary system | 5 |
|  | Eye | 4 |
|  | Musculoskeletal System | 3 |
|  | Immune system | 2 |
| AIS-C | Infections and infestations | 74 |
|  | Surgical and medical procedures | 74 |
|  | Gastrointestinal System | 44 |
|  | Pain | 43 |
|  | Psychiatric Disorders | 42 |
|  | Metabolism and nutrition | 40 |
|  | Vascular System | 37 |
|  | Respiratory System | 36 |
|  | General Disorders | 34 |
|  | Skin and subcutaneous tissue | 24 |
|  | Cardiac System | 22 |
|  | Nervous system | 21 |
|  | Blood and lymphatic system | 14 |
|  | Renal and urinary system | 8 |
|  | Eye | 7 |
|  | Ear and labyrinth | 5 |
|  | Immune system | 5 |
|  | Musculoskeletal System | 4 |
|  | Neoplasms | 2 |
|  | Reproductive system and breast | 2 |
| AIS-D | Surgical and medical procedures | 28 |
|  | Infections and infestations | 27 |
|  | Pain | 26 |
|  | Gastrointestinal System | 19 |
|  | Psychiatric Disorders | 18 |
|  | Vascular System | 11 |
|  | Metabolism and nutrition | 10 |
|  | Respiratory System | 9 |
|  | Nervous system | 7 |
|  | Blood and lymphatic system | 6 |
|  | Skin and subcutaneous tissue | 5 |
|  | General Disorders | 4 |
|  | Immune system | 2 |
|  | Renal and urinary system | 2 |
|  | Eye | 1 |
| Unknown | Infections and infestations | 71 |
|  | Surgical and medical procedures | 53 |
|  | Gastrointestinal System | 43 |
|  | Pain | 42 |
|  | Metabolism and nutrition | 34 |
|  | Psychiatric Disorders | 33 |
|  | Respiratory System | 31 |
|  | Vascular System | 29 |
|  | General Disorders | 24 |
|  | Nervous system | 20 |
|  | Cardiac System | 16 |
|  | Skin and subcutaneous tissue | 13 |
|  | Blood and lymphatic system | 12 |
|  | Renal and urinary system | 7 |
|  | Ear and labyrinth | 3 |
|  | Musculoskeletal System | 3 |
|  | Endocrine System | 1 |
|  | Eye | 1 |
|  | Immune system | 1 |

**American Spinal Injury Association Impairment Scale (AIS):* ***AIS-A****, no sensory or motor function is preserved in the sacral segments S4-5.* ***AIS-B****, sensory but no motor function is preserved below the neurological level and includes the sacral segments S4-5 (LT or PP at S4-5 or DAP), and no motor function is preserved more than three levels below the motor level on either side of the body.* ***AIS-C****, motor function is preserved at the most caudal sacral segments for voluntary anal contraction OR the patient meets the criteria for sensory incomplete status, and has some sparing of motor function more than three levels below the ipsilateral motor level on either side of the body. Less than half of key muscle functions below the single NLI have a muscle grade ≥ 3.* ***AIS-D****, motor incomplete status as defined above, with at least half (half or more) of key muscle functions below the single NLI having a muscle grade ≥ 3.* ***AIS-E****, if sensation and motor function as tested with the ISNCSCI are graded as normal in all segments, and the patient had prior deficits, then the AIS grade is E. Someone without an initial SCI does not receive an AIS grade.*
