## Supplementary material for "Pharmacological Management of Acute Spinal Cord Injury: A longitudinal multi-cohort observational study": Table e5

| Supplementary Table 5. Overview of medications administered to treat secondary complications arising from different organ systems. This data is from the Sygen clinical trial. | |
| --- | --- |
| Organ System | **Medications Administered** |
| Blood and Lymphatic System | Albumin, Atropine, Bicarbonate, Clonazepam, Cryoprecipitate, Desmopressin, Potassium Phosphate, Digoxin, Diphenhydramine, Dopamine, Enalapril Maleate, Enoxaparin, Epoetin, Famotidine, Ferrous, Ferrous Gluconate, Ferrous Sulfate, Filgrastim, Folic Acid, Fresh Frozen Plasma, Furosemide, Heparin, Hydroxyurea, Insulin, Iron, Iron Dextran, Miconazole, Nortriptyline, Phytonadione, Plasma, Platelets, Protamine, Pseudoephedrine, Red Blood Cells, Thrombin, Vitamin K, Warfarin |
| Cardiac System | Acetaminophen and Hydrocodone, Adenosine, Albumin, Aminophylline, Aspirin, Atropine, Bretylium, Calcium Chloride, Captopril, Clotrimazole, Diazepam, Digoxin, Diltiazem, Diphenhydramine, Dipyridamole, Dobutamine, Dopamine, Doxazosin, Ephedrine, Epinephrine, Epinephrine Sulfate, Esmolol, Furosemide, Glycopyrrolate, Heparin, Indomethacin, Isoproterenol, Isosorbide Dinitrate, Lidocaine, Lidocaine and Epinephrine, Lorazepam, Magnesium, Mannitol, Metoprolol, Metoprolol Hydrochlorothiazide, Midazolam, Nifedipine, Nitroglycerin, Norepinephrine, Propranolol, Scopolamine, Sodium Bicarbonate, Theophylline, Verapamil, Warfarin |
| Ear and Labyrinth | Ampicillin, Carbamide Peroxide, Epinephrine, Lidocaine, Neomycin, Neomycin Sulfate Polymyxin B Sulfate Bacitracin Zinc and Hydrocortisone, Tobramycin, Triethanolamine Polypeptide Oleate |
| Endocrine System | Dexamethasone, Insulin |
| Eye | Bacitracin, Fluorometholone, Gentamicin, Hydroxypropyl Methylcellulose, Metipranolol, Naphazoline, Neomycin Sulfate Polymyxin B Sulfate Bacitracin Zinc and Hydrocortisone, Phenylephrine, Sulfacetamide, Tobramycin, Tropicamide |
| Gastrointestinal System | Acetaminophen, Acetic Acid and Hydrocortisone, Albumin, Aluminum Hydroxide, Aluminum Magnesium, Aluminum Magnesium Simethicone, Atropine and Diphenoxylate, Bacitracin, Belladonna Alkaloids and Phenobarbital, Benzquinamide Hydrochloride, Bethanechol, Bisacodyl, Bismuth Subsalicylate, Capsaicin, Cefazolin, Cimetidine, Ciprofloxacin, Cisapride, Clotrimazole, Dexamethasone, Dextrose, Potassium Phosphate, Digoxin, Dimenhydrinate, Diphenhydramine, Docusate, Droperidol, Epinephrine, Erythromycin, Famotidine, Fentanyl, Ferrous Sulfate, Fluconazole, Fludrocortisone, Fresh Frozen Plasma, Glycerin, Glycopyrrolate, Guaifenesin Phenylephrine, Hydrocortisone, Hydroxyzine, Kaolin Pectin, Lactase, Lactobacillus, Lactulose, Lidocaine, Loperamide, Magnesium Citrate, Magnesium Hydroxide, Magnesium Oxide, Meperidine, Metoclopramide, Metronidazole, Mineral Oil, Misoprostol, Multivitamin and Supplements, Nalbuphine, Nicotine, Nitroglycerin, Nizatidine, Nystatin, Octreotide, Omeprazole, Ondansetron, Pancrelipase, Phosphate, Platelets, Polyethylene Glycol Electrolyte Solution, Potassium Acetate, Potassium Permanganate, Prochlorperazine, Prochlorperazine Maleate, Promethazine, Propantheline, Psyllium, Ranitidine, Red Blood Cells, Scopolamine, Senna, Sennosides, Simethicone, Sodium Phosphates, Sodium Polystyrene Sulfonate, Sorbitol, Sucralfate, Sulfamethoxazole, Tobramycin, Trimethobenzamide, Trimethoprim, Vancomycin, Vasopressin, Vitamin A and D |
| General Disorders | Acetaminophen, Acetaminophen and Codeine, Acetaminophen and Hydrocodone, Acetaminophen and Oxycodone, Acetaminophen and Propoxyphene, Acetazolamide, Albumin, Amikacin, Amitriptyline, Amoxicillin and Clavulanate Potassium, Ampicillin, Ampicillin and Sulbactam, Aspirin, Aztreonam, Cefazolin, Cefoperazone, Cefotaxime, Cefotetan, Ceftazidime, Ceftriaxone, Cefuroxime, Cephalexin, Chloramphenicol, Chlorhexidine, Chlorhexidine Gluconate, Chlorpromazine, Choline Magnesium Trisalicylate, Cimetidine, Ciprofloxacin, Clindamycin, Codeine, Dexamethasone, Dicloxacillin, Erythromycin, Etomidate, Fentanyl, Fluconazole, Furosemide, Gentamicin, Hydrocortisone, Hydroxyzine, Ibuprofen, Indomethacin, Ipratropium, Ketorolac, Lidocaine, Lorazepam, Mecamylamine, Meclizine, Meperidine, Methylprednisolone, Metronidazole, Midazolam, Morphine, Nafcillin, Naproxen, Ofloxacin, Oxacillin, Penicillin, Piperacillin, Piperacillin and Tazobactam, Piroxicam, Promethazine, Propoxyphene, Pseudoephedrine, Saline, Scopolamine, Sodium Chloride, Sulfamethoxazole, Ticarcillin, Tobramycin, Trimethoprim, Vancomycin |
| Immune System | Acetazolamide, Astemizole, Cimetidine, Diphenhydramine, Furosemide, Hydrocortisone, Hydroxyzine, Ketotifen, Methylprednisolone Sodium Succinate, Prednisone, Ranitidine, Red Blood Cells |
| Infections and Infestations | Acetazolamide, Acetic Acid, Acetylcysteine, Acyclovir, Albuterol, Alcohol, Aluminum Magnesium, Amikacin, Aminophylline, Amitriptyline, Amoxicillin, Amoxicillin and Clavulanate Potassium, Amphotericin B, Ampicillin, Ampicillin and Sulbactam, Antipyrine Benzocaine Otic, Antipyrine Benzocaine U-Polycosanol, Atropine, Azithromycin, Aztreonam, Bacitracin, Baclofen, Beclomethasone, Betamethasone and Clotrimazole, Bismuth Subsalicylate, Brompheniramine Pseudoephedrine, Butoconazole Nitrate, Carbenicillin, Carbenicillin Indanyl, Cefaclor, Cefadroxil, Cefamandole, Cefazolin, Cefoperazone, Cefotaxime, Cefotetan, Cefoxitin, Cefpodoxime, Ceftazidime, Ceftizoxime, Ceftriaxone, Cefuroxime, Cephalexin, Cephalosporin, Cephapirin, Cephradine, Chlordiazepoxide, Chlorhexidine, Chlorpheniramine, Cimetidine, Ciprofloxacin, Cisapride, Clarithromycin, Clindamycin, Clotrimazole, Cloxacillin, Corticosteroids, Dexamethasone, Diclofenac, Dicloxacillin, Diphenhydramine, Diptheria Pertussis and Tetanus Vaccine, Doxycycline, Epinephrine, Erythromycin, Etomidate, Fentanyl, Fluconazole, Fluoxetine, Furosemide, Gentamicin, Glycopyrrolate, Guaifenesin, Guaifenesin Phenylephrine, Heparin, Hydrocortisone, Ibuprofen, Imipenem, Imipenem and Cilastatin, Indomethacin, Influenza Vaccine, Ipratropium, Ketoconazole, Ketorolac, Lactobacillus, Lidocaine, Meperidine, Meropenem, Methenamine, Methylprednisolone, Methylprednisolone Sodium Succinate, Metronidazole, Mezlocillin, Miconazole, Midazolam, Minocycline, Morphine, Mupirocin, Nabumetone, Nafcillin, Naproxen, Neomycin, Neomycin Sulfate Polymyxin B Sulfate Bacitracin Zinc and Hydrocortisone, Neostigmine, Nitrofurantoin, Norepinephrine, Norfloxacin, Nystatin, Nystatin Triamcinolone, Ofloxacin, Oxacillin, Oxybutynin, Oxymetazoline, Pancrelipase, Pancuronium Bromide, Penicillin, Permethrin, Phenazopyridine, Phenol, Phenytoin, Piperacillin, Piperacillin and Tazobactam, Pneumococcal Vaccine, Potassium Chloride, Povidone Iodine, Prednisolone Acetate, Prednisone, Promazine, Pseudoephedrine, Ranitidine, Rifampin, Saline, Scopolamine, Silver Sulfadiazine, Sodium Chloride, Sulfacetamide, Sulfamethoxazole, Sulindac, Terazosin, Terconazole, Terfenadine, Tetanus Vaccine, Tetracycline, Ticarcillin, Tobramycin, Tobramycin and Dexamethasone, Tolnaftate, Tolterodine, Trimethoprim, Vancomycin |
| Metabolism and Nutrition | Acetate, Acetazolamide, Albumin, Aluminum Hydroxide, Aluminum Magnesium Simethicone, Amino Acids, Bicarbonate, Bisacodyl, Calcium, Calcium Carbide, Calcium Carbonate, Calcium Carbonate and Magnesium Chloride, Calcium Chloride, Calcium Citrate, Calcium Gluceptate, Calcium Gluconate, Calcium Gluconate and Sodium Phosphates, Carbonic Acid, Chloride, Cholestyramine, Cimetidine, Cisatracurium, Desmopressin, Dextrose, Magnesium Sulfate, Potassium Chloride, Sodium Acetate, Dimenhydrinate, Etidronate, Famotidine, Ferrous Gluconate, Ferrous Sulfate, Fludrocortisone, Fluoxetine, Folic Acid, Fresh Frozen Plasma, Furosemide, Glipizide, Glucose, Glyburide, Insulin, Ipratropium, Iron Dextran, Iron Vitamin C, Isoflurane, Potassium Chloride, Lactase, Lactobacillus, Lactulose, Lipids, Magnesium, Magnesium Gluconate, Magnesium Hydroxide, Magnesium Oxide, M Megestrol Acetate, Methylphenidate, Metoclopramide, Morphine, Multivitamin and Supplements, Pamidronate, Phosphate, Phytonadione, Platelets, Polyethylene Glycol Electrolyte Solution, Potassium, Potassium Acetate, Potassium Phosphate, Potassium Phosphate and Sodium Phosphates, Ranitidine, Saline, Selenium, Sodium, Sodium Bicarbonate, Sodium Bisphosphate and Sodium Phosphates, Sodium Chloride, Sodium Phosphates, Sodium Phosphates, Sodium Polystyrene Sulfonate, Sucralfate, Tolbutamide, Urea, Vancomycin, Vitamin B, Vitamin B1, Vitamin C, Vitamin D3, Vitamin K3, Zinc |
| Musculoskeletal System | Alendronate, Dantrolene, Diazepam, Etidronate, Indomethacin, Naproxen, Pancuronium Bromide, Prazosin, Quinine Sulfate, Vecuronium Bromide |
| Neoplasms | Allopurinol, Hydroxyurea |
| Nervous System | Acetaminophen and Methocarbamol, Acetaminophen and Oxycodone, Acetazolamide, Amitriptyline, Baclofen, Belladonna Alkaloids Opium, Benztropine, Calcium Gluceptate, Capsaicin, Carbamazepine, Carisoprodol, Cefamandole, Chlorzoxazone, Choline Magnesium Trisalicylate, Clonazepam, Clonidine, Cyclobenzaprine, Dantrolene, Desipramine, Dexamethasone, Diazepam, Divalproex Sodium, Dopamine, Ephedrine, Etidronate, Famotidine, Fludrocortisone, Furosemide, Gabapentin, Haloperidol, Ibuprofen, Insulin, Ketorolac, Levodopa, Lorazepam, Magnesium Sulfate, Mannitol, Methocarbamol, Methylprednisolone, Methylprednisolone Sodium Succinate, Morphine, Naproxen, Nifedipine, Nitrofurantoin, Nortriptyline, Oxybutynin, Phenazopyridine, Phenobarbital, Phenoxybenzamine, Phenylephrine, Phenytoin, Potassium Iodide, Terazosin, Tramadol, Xylometazoline Hydrogen chloride |
| Pain | Acetaminophen, Acetaminophen and Codeine, Acetaminophen and Hydrocodone, Acetaminophen and Oxycodone, Acetaminophen and Propoxyphene, Aluminum Magnesium Simethicone, Amitriptyline, Aspirin, Aspirin and Oxycodone, Aspirin Butalbital and Caffeine, Bacitracin, Baclofen, Benzocaine, Benzocaine Menthol, Bupivacaine, Buprenorphine, Butamben Tetracaine Benzocaine, Butorphanol Tartrate, Capsaicin, Carbamazepine, Carbamide Peroxide, Cocaine, Codeine, Codeine Phosphate, Codeine Sulfate, Cyclobenzaprine, Desipramine, Dexamethasone, Midazolam, Diazepam, Diclofenac, Enalapril Maleate, Etodolac, Fentanyl, Fludrocortisone, Gabapentin, Gentamicin, Haloperidol, Hydrocodone, Hydromorphone, Hydroxyzine, Ibuprofen, Indomethacin, Ketoprofen, Ketorolac, Levorphanol, Lidocaine, Lorazepam, Magnesium Sulfate, Meperidine, Meperidine and Hydroxyzine, Methadone, Methadone and Acetaminophen, Methocarbamol, Methyl Salicylate, Methylprednisolone, Metoclopramide, Morphine, Morphine Sulfate, Nabumetone, Nalbuphine, Naloxone, Naproxen, Neomycin Sulfate Polymyxin B Sulfate Bacitracin Zinc and Hydrocortisone, Norfloxacin, Nortriptyline, Nystatin, Omeprazole, Oxaprozin, Oxycodone, Pentobarbital, Phenobarbital, Phenol, Potassium Chloride, Promethazine, Propoxyphene, Tramadol, Trazodone, Triazolam, Triethanolamine Polypeptide Oleate, Trolamine Salicylate, Zinc Sulfate Monohydrate |
| Psychiatric Disorders | Acetaminophen and Hydrocodone, Alprazolam, Amitriptyline, Benztropine, Bisacodyl, Bupropion, Buspirone, Butamben Tetracaine Benzocaine, Butorphanol Tartrate, Chloral Hydrate, Chlordiazepoxide, Chlorpromazine, Clonazepam, Clonidine, Clorazepate, Codeine, Desipramine, Dextromethorphan Guaifenesin and Pseudoephedrine, Fentanyl, Diazepam, Dimenhydrinate, Diphenhydramine, Doxepin, Droperidol, Ephedrine, Estazolam, Ethanol, Fluoxetine, Fluphenazine, Flurazepam, Haloperidol, Hydromorphone, Hydroxyzine, Imipenem, Imipramine, Imipramine Pamoate, Levomepromazine, Lithium, Lorazepam, Meperidine, Methocarbamol, Methylphenidate, Metoclopramide, Midazolam, Morphine, Multivitamin and Supplements, Naloxone, Nefazodone, Neostigmine, Nicotine, Nortriptyline, Oxazepam, Paroxetine, Pentobarbital, Phenobarbital, Phenytoin, Promethazine, Propofol, Pseudoephedrine, Quazepam, Ranitidine, Risperidone, Sertraline, Terazosin, Thioridazine, Trazodone, Triazolam, Trifluoperazine , Vecuronium Bromide, Venlafaxine, Vitamin B1, Zolpidem |
| Renal and Urinary System | Acetazolamide, Aminocaproic Acid, Amitriptyline, Amphotericin B, Bethanechol, Bumetanide, Cefazolin, Chlorothiazide, Ciprofloxacin, Desmopressin, Dopamine, Furosemide, Gentamicin, Levodopa, Methenamine, Metoclopramide, Metolazone, Metronidazole, Mezlocillin, Neostigmine, Oxybutynin, Propantheline, Ranitidine, Saline, Sennosides, Sodium Bicarbonate, Terazosin, Tolterodine |
| Reproductive System and Breast | Conjugated Estrogens, Estrogen, Progesterone |
| Respiratory System | Acetaminophen Dexbrompheniramine Pseudoephedrine, Acetylcysteine, Acetylcysteine and Albuterol, Albumin, Albuterol, Aminophylline, Ammonium Chloride, Bacitracin, Baclofen, Beclomethasone, Benzocaine Menthol, Bicarbonate, Brompheniramine, Brompheniramine Dextromethorphan and Pseudoephedrine, Brompheniramine Pseudoephedrine, Cefaclor, Cefazolin, Cefotaxime, Cefotetan, Ceftazidime, Ceftriaxone, Cefuroxime, Cephalexin, Chlorpheniramine, Chlorpheniramine Methscopolamine Phenylephrine, Chlorpromazine, Clarithromycin, Clindamycin, Cromolyn Sodium, Dexamethasone, Diazepam, Diphenhydramine, Docusate, Ephedrine, Epinephrine, Epinephrine, Epinephrine Sulfate, Erythromycin, Fentanyl, Fluconazole, Furosemide, Gentamicin, Glycopyrrolate, Guaifenesin, Guaifenesin and Theophylline, Guaifenesin Phenylephrine, Guaifenesin Potassium Guaiacolsulfonate, Haloperidol, Hydrocortisone, Imipenem, Ipratropium, Ipratropium Bromide, Isoflurane, Ketotifen, Lidocaine, Lorazepam, Metaproterenol, Methylprednisolone, Methylprednisolone Sodium Succinate, Metoclopramide, Metocurine Iodide, Metolazone, Metronidazole, Mezlocillin, Midazolam, Morphine, Naloxone, Nitroglycerin, Norepinephrine, Norfloxacin, Ofloxacin, Oxymetazoline, Phenylephrine, Phenylpropanolamine, Piperacillin, Pirbuterol, Potassium Iodide, Prednisone, Prochlorperazine, Pseudoephedrine, Pseudoephedrine Triprolidine, Red Blood Cells, Rifampin, Saline, Salmeterol , Sodium Bicarbonate, Sodium Chloride, Sulfamethoxazole, Terbutaline, Terfenadine, Theophylline, Ticarcillin, Tobramycin, Triamcinolone, Trimethoprim, Vancomycin, Vecuronium Bromide, Xylometazoline Hydrogen chloride |
| Skin and Subcutaneous Tissue | Aluminum Magnesium, Ammonium Lactate, Ampicillin and Sulbactam, Bacitracin, Benzoyl Peroxide, Betamethasone and Clotrimazole, Cefazolin, Cefotaxime, Ceftazidime, Cephalexin, Chlorhexidine Gluconate, Ciprofloxacin, Clindamycin, Clotrimazole, Collagenase, Cyproheptadine, Desipramine, Dexamethasone, Dicloxacillin, Diphenhydramine, Erythromycin, Fluconazole, Framycetin, Fusidic Acid, Gentamicin, Griseofulvin, Hydrocortisone, Hydrogen Peroxide, Hydroxyzine, Ketoconazole, Lidocaine, Lindane, Mafenide, Merbromin, Miconazole, Mineral Oil, Minoxidil, Mupirocin, Nystatin, Pancuronium Bromide, Penicillin, Podofilox, Potassium Chloride, Potassium Permanganate, Salicylic Acid, Saline, Selenium Sulfide, Silver Sulfadiazine, Sodium Chloride, Sodium Hypochlorite, Sulfacetamide Sodium and Urea, Terbinafine Hydrochloride, Terconazole, Tetracycline, Tobramycin and Dexamethasone, Triamcinolone, Vancomycin, Vitamin C, Zinc, Zinc Oxide, Zinc Sulfate Monohydrate |
| Surgical and Medical Procedures | Acetaminophen and Codeine, Acetaminophen and Oxycodone, Albumin, Albuterol, Amikacin, Aminophylline, Ampicillin, Ampicillin and Sulbactam, Atracurium, Atropine, Bacitracin, Benzocaine, Benzocaine Menthol, Benzoyl Peroxide, Bupivacaine, Bupivacaine Hydrochloride and Epinephrine Bitartrate, Butamben Tetracaine Benzocaine, Butorphanol, Butorphanol Tartrate, Cefazolin, Cefotetan, Ceftazidime, Ceftizoxime, Ceftriaxone, Cefuroxime, Cephalexin, Chloral Hydrate, Cimetidine, Ciprofloxacin, Cisapride, Cisatracurium, Clonazepam, Cloxacillin, Cocaine, Codeine, Cyclobenzaprine, Desflurane, Dexamethasone, Fentanyl, Heparin, Vecuronium Bromide, Dezocine, Diatrizoate Meglumine and Diatrizoate Sodium, Diazepam, Dibucaine, Dimenhydrinate, Diphenhydramine, DOM, Droperidol, Enoxaparin, Ephedrine, Epinephrine, Epinephrine Sulfate, Etidronate, Etomidate, Famotidine, Fluconazole, Flumazenil, Flurazepam, Furosemide, Gelatin, Gentamicin, Glucagon, Glycopyrrolate, Haloperidol, Hydralazine, Hydrocortisone, Hydromorphone, Hydroxyzine, Iohexol, Iothalamate Meglumine, Isoflurane, Potassium Chloride, Ketamine, Ketorolac, Labetalol, Levodopa, Lidocaine, Lidocaine and Epinephrine, Lidocaine and Prilocaine, Lorazepam, Magnesium Citrate, Magnesium Sulfate, Mannitol, Meperidine, Metacyclin, Methocarbamol, Methylprednisolone, Methylprednisolone Sodium Succinate, Metoclopramide, Metocurine Iodide, Metronidazole, Metubine, Midazolam, Mivacurium Chloride, Morphine, Nafcillin, Naloxone, Neostigmine, Nitroglycerin, Nitrous Oxide, Norepinephrine, Ofloxacin, Oxacillin, Pancrelipase, Pancuronium Bromide, Penicillin, Pentobarbital, Phenylephrine, Pipecuronium, Piperacillin, Platelets, Polyethylene Glycol Electrolyte Solution, Potassium Chloride, Povidone Iodine, Prilocaine, Propofol, Ranitidine, Red Blood Cells, Rocuronium, Saline, Sevoflurane, Sodium Bicarbonate, Sodium Chloride, Sodium Chloride Sodium Gluconate Sodium Acetate Potassium Chloride and Magnesium Chloride, Succinylcholine, Sufentanil, Sulfamethoxazole, Tetracycline, Thiopental, Thrombin, Tobramycin, Trimethoprim, Urokinase, Vancomycin, Vitamin B |
| Vascular System | Acetazolamide, Albumin, Amlodipine, Aspirin, Atenolol, Atropine, Captopril, Chlordiazepoxide, Ciprofloxacin, Clonidine, Dalteparin, Desmopressin, Dexamethasone, Dextrose, Dobutamine, Dopamine, Heparin, Potassium Chloride, Diltiazem, Enalapril, Enalapril Maleate, Enoxaparin, Ephedrine, Epinephrine, Epinephrine Sulfate, Esmolol, Fluconazole, Fludrocortisone, Fresh Frozen Plasma, Furosemide, Hydralazine, Hydrochlorothiazide, Hydrocortisone Acetate, Hydroxyzine, Insulin, Isoproterenol, Labetalol, Lidocaine, Lisinopril, Magnesium Sulfate, Mannitol, Metolazone, Metoprolol, Metoprolol Hydrochlorothiazide, Midazolam, Morphine, Nifedipine, Nitroglycerin, Nizatidine, Norepinephrine, Nortriptyline, Phenoxybenzamine, Phenylephrine, Plasma, Platelets, Potassium, Potassium Phosphate, Pramoxine Phenylephrine Glycerin Petrolatum, Propranolol, Pseudoephedrine, Quinapril, Red Blood Cells, Saline, Sodium Bicarbonate, Sodium Chloride, Sodium Chloride Sodium Gluconate Sodium Acetate Potassium Chloride and Magnesium Chloride, Urea, Vasopressin, Vitamin K, Warfarin, Zinc Sulfate Monohydrate |
