## Supplementary material for "Pharmacological Management of Acute Spinal Cord Injury: A longitudinal multi-cohort observational study": Table e6

| **Supplementary Table 6. Ability of drugs to pass the blood brain barrier (BBB).** The probabilities of passing the blood brain barrier (or not) were extracted from drugbank.ca or literature. | | | |
| --- | --- | --- | --- |
| **Drug** | **BBB** | **Probability** | **Source** |
| Acarbose | No | 0.9723 | drugbank.ca |
| Acetylcysteine | No |  | drugbank.ca |
| Adalimumab | No |  | drugbank.ca |
| Albuterol | No | 0.9659 | drugbank.ca |
| Alpha d galactosidase | No |  | drugbank.ca |
| Amikacin | No | 0.9659 | drugbank.ca |
| Amlodipine | No | 0.7744 | drugbank.ca |
| Amoxicillin | No | 0.9967 | drugbank.ca |
| Amoxicillin clavulanate | No | 0.9967 | drugbank.ca |
| Amoxicillin clavulanate potassium | No | 0.9967 | drugbank.ca |
| Amphotericin b | No | 0.9659 | drugbank.ca |
| Ampicillin | No | 0.9961 | drugbank.ca |
| Ampicillin sulbactam | No |  | drugbank.ca |
| Anidulafungin | No | 0.9833 | drugbank.ca |
| Argatroban | No | 0.8933 | drugbank.ca |
| Atenolol | No | 0.9505 | drugbank.ca |
| Atorvastatin | No | 0.7825 | drugbank.ca |
| Azithromycin | No | 0.9739 | drugbank.ca |
| Aztreonam | No | 0.9657 | drugbank.ca |
| Bacitracin | No | 0.9943 | drugbank.ca |
| Bacitracin neomycin polymyxin b | No | 0.9943 | drugbank.ca |
| Bacitracin polymyxin b | No | 1.9943 | drugbank.ca |
| Benazepril | No | 0.7082 | drugbank.ca |
| Bisoprolol | No | 0.9077 | drugbank.ca |
| Bromocriptine | No | 0.9845 | drugbank.ca |
| Bumetadine | No | 0.6636 | drugbank.ca |
| Capsaicin | No |  | PMID:3696465 |
| Carbenicillin | No | 0.9954 | drugbank.ca |
| Carbenicillin indanyl | No | 0.9954 | drugbank.ca |
| Carbidopa | No | 0.9659 | drugbank.ca |
| Carvedilol | No | 0.9332 | drugbank.ca |
| Caspofungin | No | 0.96 | drugbank.ca |
| Cefaclor | No | 0.9817 | drugbank.ca |
| Cefadroxil | No | 0.9966 | drugbank.ca |
| Cefamandole | No | 0.9918 | drugbank.ca |
| Cefazolin | No | 0.987 | drugbank.ca |
| Cefepime | No | 0.9332 | drugbank.ca |
| Cefoperazone | No | 0.9886 | drugbank.ca |
| Cefotaxime | No | 0.9877 | drugbank.ca |
| Cefotetan | No | 0.9903 | drugbank.ca |
| Cefoxitin | No | 0.9952 | drugbank.ca |
| Cefpodoxime | No | 0.984 | drugbank.ca |
| Ceftazidime | No | 0.9857 | drugbank.ca |
| Ceftizoxime | No | 0.9823 | drugbank.ca |
| Ceftriaxone | No | 0.9748 | drugbank.ca |
| Cefuroxime | No | 0.9863 | drugbank.ca |
| Cephalexin | No | 0.996 | drugbank.ca |
| Cephalosporin | No | 0.9961 | drugbank.ca |
| Cephapirin | No | 0.9971 | drugbank.ca |
| Cephradine | No | 0.9942 | drugbank.ca |
| Chlorothiazide | No | 0.9506 | drugbank.ca |
| Choline magnesium | No | 0.8993 | drugbank.ca |
| Choline magnesium trisalicylate | No | 0.8993 | drugbank.ca |
| Cimetidine | No | 0.6782 | drugbank.ca |
| Clarithromycin | No | 0.987 | drugbank.ca |
| Clindamycin | No | 0.9697 | drugbank.ca |
| Cloxacillin | No | 0.9903 | drugbank.ca |
| Colistimethate | No | 0.8404 | drugbank.ca |
| Demeclocycline | No | 0.9659 | drugbank.ca |
| Desmopressin | No | 0.8866 | drugbank.ca |
| Dicloxacillin | No | 0.9903 | drugbank.ca |
| Digoxin | No | 0.7241 | drugbank.ca |
| Dipyridamole | No | 0.6246 | drugbank.ca |
| Dobutamine | No | 0.7448 | drugbank.ca |
| Dopamine | No | 0.8414 | drugbank.ca |
| Doripenem | No | 0.9633 | drugbank.ca |
| Enalapril | No | 0.9659 | drugbank.ca |
| Enalapril hydrochlorothiazide | No | 0.9659 | drugbank.ca |
| Enalapril maleate | No | 0.9659 | drugbank.ca |
| Enoxaparin | No | 0.8366 | drugbank.ca |
| Epinephrine | No | 0.966 | drugbank.ca |
| Epinephrine lidocaine | No | 0.966 | drugbank.ca |
| Epinephrine sulfate | No | 0.966 | drugbank.ca |
| Ergotamine caffeine | No | 0.9644 | drugbank.ca |
| Ertapenem | No | 0.9811 | drugbank.ca |
| Erythromycin | No | 0.9889 | drugbank.ca |
| Esmolol | No | 0.9645 | drugbank.ca |
| Esomeprazole | No | 0.6326 | drugbank.ca |
| Felodipine | No | 0.83 | drugbank.ca |
| Fexofenadine | No | 0.8574 | drugbank.ca |
| Fondaparinux | No | 0.8953 | drugbank.ca |
| Fosinopril | No | 0.667 | drugbank.ca |
| Furosemide | No | 0.8833 | drugbank.ca |
| Gatifloxacin | No | 0.9869 | drugbank.ca |
| Gentamicin | No | 0.9826 | drugbank.ca |
| Glucosamine | No | 0.9143 | drugbank.ca |
| Glucosamine chondroitin | No | 0.9143 | drugbank.ca |
| Glucose | No | 0.5569 | drugbank.ca |
| Glyburide | No | 0.6707 | drugbank.ca |
| Guaifenesin | No | 0.8631 | drugbank.ca |
| Guaifenesin potassium guaiacolsulfonate | No |  | drugbank.ca |
| Hydrochlorothiazide triamterene | No | 0.9659 | drugbank.ca |
| Ibandronate sodium | No | 0.5229 | drugbank.ca |
| Imipenem cilastatin | No | 0.9711 | drugbank.ca |
| Iohexol | No | 0.5082 | drugbank.ca |
| Labetalol | No | 0.8313 | drugbank.ca |
| Levodopa | No | 0.9264 | drugbank.ca |
| Liothyronine | No | 0.6886 | drugbank.ca |
| Lisinopril | No | 0.8815 | drugbank.ca |
| Losartan | No | 0.7812 | drugbank.ca |
| Losartan hydrochlorothiazide | No |  | drugbank.ca |
| Mannitol | No | 0.5997 | drugbank.ca |
| Meloxicam | No | 0.9811 | drugbank.ca |
| Meropenem | No | 0.9901 | drugbank.ca |
| Metaproterenol | No | 0.951 | drugbank.ca |
| Methotrexate | No | 0.9467 | drugbank.ca |
| Methyldopa | No | 0.9276 | drugbank.ca |
| Metipranolol | No | 0.9906 | drugbank.ca |
| Metoprolol | No | 0.8426 | drugbank.ca |
| Mezlocillin | No | 0.9802 | drugbank.ca |
| Micafungin | No | 0.8524 | drugbank.ca |
| Midodrine | No | 0.856 | drugbank.ca |
| Minocycline | No | 0.9783 | drugbank.ca |
| Moxifloxacin | No | 0.9597 | drugbank.ca |
| Mupirocin | No | 0.5988 | drugbank.ca |
| Nafcillin | No | 0.9938 | drugbank.ca |
| Neomycin | No | 0.9659 | drugbank.ca |
| Nicardipine | No | 0.9549 | drugbank.ca |
| Nifedipine | No | 0.9536 | drugbank.ca |
| Nisoldipine | No | 0.947 | drugbank.ca |
| Norfloxacin | No | 0.9824 | drugbank.ca |
| Octreotide | No | 0.8738 | drugbank.ca |
| Ofloxacin | No | 0.9659 | drugbank.ca |
| Olmesartan medoxomil | No | 0.8719 | drugbank.ca |
| Omeprazole | No | 0.6326 | drugbank.ca |
| Oseltamivir | No | 0.9535 | drugbank.ca |
| Oxacillin | No | 0.9923 | drugbank.ca |
| Oxymetazoline | No | 0.5388 | drugbank.ca |
| Pamipril | No | 0.5777 | drugbank.ca |
| Pancrelipase | No |  | drugbank.ca |
| Penicillin | No | 0.9954 | drugbank.ca |
| Phenylephrine | No | 0.9115 | drugbank.ca |
| Pimecrolimus | No | 0.9554 | drugbank.ca |
| Piperacillin | No | 0.9958 | drugbank.ca |
| Piperacillin tazobactam | No |  | drugbank.ca |
| Piroxicam | No | 0.9659 | drugbank.ca |
| Podofilox | No | 0.5388 | drugbank.ca |
| Posaconazole | No | 0.5499 | drugbank.ca |
| Propranolol | No | 0.9031 | drugbank.ca |
| Quinapril | No | 0.9409 | drugbank.ca |
| Ramipril | No | 0.8726 | drugbank.ca |
| Ranitidine | No | 0.8783 | drugbank.ca |
| Ranolazine | No | 0.5334 | drugbank.ca |
| Repaglinide | No | 0.7101 | drugbank.ca |
| Rifampin | No | 0.974 | drugbank.ca |
| Rifaximin | No | 0.973 | drugbank.ca |
| Ringers lactate | No |  | drugbank.ca |
| Rosuvastatin | No | 0.6815 | drugbank.ca |
| Simethicone | No |  | drugbank.ca |
| Sorbitol | No | 0.5997 | drugbank.ca |
| Terbutaline | No | 0.9355 | drugbank.ca |
| Tetracycline | No | 0.9841 | drugbank.ca |
| Ticarcillin | No | 0.9921 | drugbank.ca |
| Ticarcillin potassium clavulanate | No | 0.9921 | drugbank.ca |
| Tigecycline | No | 0.9836 | drugbank.ca |
| Tobramycin | No | 0.9794 | drugbank.ca |
| Tobramycin dexamethasone | No |  | drugbank.ca |
| Valsartan | No | 0.8032 | drugbank.ca |
| Vancomycin | No | 0.991 | drugbank.ca |
| Vitamin b12 | No | 0.7447 | drugbank.ca |
| Acetylcysteine albuterol | Unknown |  |  |
| Acetylsalicylic acid butalbital caffeine | Unknown |  |  |
| Aliskiren | Unknown |  |  |
| Allantoin camphor phenol | Unknown |  |  |
| Aluminum magnesium simethicone | Unknown |  |  |
| Amylase lipase protease | Unknown |  |  |
| Atracurium | Unknown |  |  |
| Barium sulfate | Unknown |  |  |
| Becaplermin | Unknown |  |  |
| Benzoyl peroxide | Unknown |  |  |
| Betamethasone clotrimazole | Unknown |  |  |
| Botulinum toxin | Unknown |  |  |
| Butamben tetracaine benzocaine | Unknown |  |  |
| Calamine phenolate | Unknown |  |  |
| Calcium carbonate | Unknown |  |  |
| Calcium carbonate magnesium chloride | Unknown |  |  |
| Calcium citrate | Unknown |  |  |
| Calcium gluconate | Unknown |  |  |
| Calcium gluconate sodium phosphate | Unknown |  |  |
| Calcium polycarbophil | Unknown |  |  |
| Calcium simethicone | Unknown |  |  |
| Camphor menthol | Unknown |  |  |
| Camphor phenol | Unknown |  |  |
| Camphor pramoxine zinc | Unknown |  |  |
| Candesartan | Unknown |  |  |
| Carboxymethylcellulose sodium | Unknown |  |  |
| Cetirizine pseudoephedrine | Unknown |  |  |
| Chloraseptic | Unknown |  |  |
| Chloride | Unknown |  |  |
| Cholestyramine | Unknown |  |  |
| Ciprofloxacin hydrocortisone | Unknown |  |  |
| Citric acid sodium citrate | Unknown |  |  |
| Clobetasol | Unknown |  |  |
| Collagenase | Unknown |  |  |
| Corticosteroids | Unknown |  |  |
| Cortisone | Unknown |  |  |
| Cosyntropin | Unknown |  |  |
| Cryoprecipitate | Unknown |  |  |
| Cyto-zyme | Unknown |  |  |
| Dalteparin | Unknown |  |  |
| Daptomycin | Unknown |  |  |
| Darbepoetin | Unknown |  |  |
| Diatrizoate meglumine diatrizoate sodium | Unknown |  |  |
| Diphenoxylate atropine | Unknown |  |  |
| Diptheria pertussis tetanus vaccine | Unknown |  |  |
| Docusate | Unknown |  |  |
| Docusate benzocaine | Unknown |  |  |
| Docusate glycerine | Unknown |  |  |
| Docusate senna | Unknown |  |  |
| Dornase alfa | Unknown |  |  |
| Exenatide | Unknown |  |  |
| Ferrous sulfate | Unknown |  |  |
| Fluocinolone | Unknown |  |  |
| Fluticasone | Unknown |  |  |
| Fluticasone salmeterol | Unknown |  |  |
| Framycetin | Unknown |  |  |
| Gelatin | Unknown |  |  |
| Glucagon | Unknown |  |  |
| Glucoscan | Unknown |  |  |
| Glycerine | Unknown |  |  |
| Guaifenesin codeine | Unknown |  |  |
| Guaifenesin dextromethorphan | Unknown |  |  |
| Guaifenesin phenylephrine | Unknown |  |  |
| Guaifenesin theophylline | Unknown |  |  |
| Hetastarch | Unknown |  |  |
| Hydrocodone ibuprofen | Unknown |  |  |
| Hydrocortisone acetate | Unknown |  |  |
| Hydrogen peroxide | Unknown |  |  |
| Hydroxypropyl methylcellulose | Unknown |  |  |
| Immune globulin | Unknown |  |  |
| Influenza vaccine | Unknown |  |  |
| Insulin | Unknown |  |  |
| Iopamidol | Unknown |  |  |
| Iothalamate meglumine | Unknown |  |  |
| Ipratropium albuterol | Unknown |  |  |
| Iron dextran | Unknown |  |  |
| Iron vitamin c | Unknown |  |  |
| Isometheptene dichloralphenazone acetaminophen | Unknown |  |  |
| Lactase | Unknown |  |  |
| Lactobacillus | Unknown |  |  |
| Lanthanum carbonate | Unknown |  |  |
| Leuprolide acetate | Unknown |  |  |
| Levalbuterol | Unknown |  |  |
| Levofloxacin | Unknown |  |  |
| Lipids | Unknown |  |  |
| Mafenide | Unknown |  |  |
| Magnesium | Unknown |  |  |
| Magnesium chloride | Unknown |  |  |
| Magnesium gluconate | Unknown |  |  |
| Magnesium hydroxide | Unknown |  |  |
| Meperidine hydroxyzine | Unknown |  |  |
| Meperidine phenylephrine | Unknown |  |  |
| Merbromin | Unknown |  |  |
| Methenamine | Unknown |  |  |
| Methyl salicylate | Unknown |  |  |
| Methylcellulose | Unknown |  |  |
| Methylprednisolone sodium succinate | Unknown |  |  |
| Metoprolol hydrochlorothiazide | Unknown |  |  |
| Metubine | Unknown |  |  |
| Multivitamin folate | Unknown |  |  |
| Multivitamin iron | Unknown |  |  |
| Multivitamin minerals | Unknown |  |  |
| Multivitamin zinc | Unknown |  |  |
| Multivitamins | Unknown |  |  |
| Mycobacterium tuberculosis extract | Unknown |  |  |
| Nutritional supplement | Unknown |  |  |
| Omega 3 | Unknown |  |  |
| Oxychlorosene powder | Unknown |  |  |
| Phenothiazines | Unknown |  |  |
| Phosphate | Unknown |  |  |
| Plasma | Unknown |  |  |
| Platelets | Unknown |  |  |
| Pneumococcal vaccine | Unknown |  |  |
| Polycarbophil | Unknown |  |  |
| Polyethylene glycol | Unknown |  |  |
| Polyethylene glycol electrolyte solution | Unknown |  |  |
| Potassium | Unknown |  |  |
| Potassium acetate | Unknown |  |  |
| Potassium permanganate | Unknown |  |  |
| Potassium phosphate | Unknown |  |  |
| Povidone iodine | Unknown |  |  |
| Pramipexole | Unknown |  |  |
| Pramoxine | Unknown |  |  |
| Pramoxine phenylephrine glycerin petrolatum | Unknown |  |  |
| Pravastatin | Unknown |  |  |
| Protamine | Unknown |  |  |
| Pseudoephedrine cetirizine | Unknown |  |  |
| Pseudoephedrine hydrochlorothiazide | Unknown |  |  |
| Pseudoephedrine loratadine | Unknown |  |  |
| Pseudoephedrine triprolidine | Unknown |  |  |
| Psyllium | Unknown |  |  |
| Red blood cells | Unknown |  |  |
| Salicylic acid sulfur | Unknown |  |  |
| Saline | Unknown |  |  |
| Selenium | Unknown |  |  |
| Sennosides | Unknown |  |  |
| Silver nitrate | Unknown |  |  |
| Sodium | Unknown |  |  |
| Sodium acetate | Unknown |  |  |
| Sodium chloride | Unknown |  |  |
| Sodium citrate | Unknown |  |  |
| Sodium ferric gluconate complex | Unknown |  |  |
| Sodium hypochlorite | Unknown |  |  |
| Sodium phosphate | Unknown |  |  |
| Sodium polystyrene sulfonate | Unknown |  |  |
| Teriparatide | Unknown |  |  |
| Tetanus vaccine | Unknown |  |  |
| Tinzaparin | Unknown |  |  |
| Triethanolamine polypeptide | Unknown |  |  |
| Trimethoprim polymyxin b | Unknown |  |  |
| Trimethoprim sulfamethoxazole | Unknown |  |  |
| Trolamine salicylate | Unknown |  |  |
| Urea | Unknown |  |  |
| Urokinase | Unknown |  |  |
| Vasopressin | Unknown |  |  |
| Acetaminophen | Yes | 0.9544 | drugbank.ca |
| Acetaminophen aspirin | Yes |  |  |
| Acetaminophen aspirin caffeine | Yes |  |  |
| Acetaminophen butalbital caffeine | Yes |  |  |
| Acetaminophen caffeine | Yes |  |  |
| Acetaminophen codeine | Yes |  |  |
| Acetaminophen dexbrompheniramine pseudoephedrine | Yes |  |  |
| Acetaminophen dichloralphenazone isometheptene | Yes |  |  |
| Acetaminophen hydrocodone | Yes |  |  |
| Acetaminophen isometheptene | Yes |  |  |
| Acetaminophen methadone | Yes |  |  |
| Acetaminophen oxycodone | Yes |  |  |
| Acetaminophen propoxyphene | Yes |  |  |
| Acetate | Yes |  | PMID 19393008 |
| Acetazolamide | Yes | 0.9382 | drugbank.ca |
| Acetic acid | Yes | 0.9742 | drugbank.ca |
| Acetic acid hydrocortisone | Yes | 0.9742 | drugbank.ca |
| Acetylsalicylic acid | Yes | 0.9376 | drugbank.ca |
| Acetylsalicylic acid oxycodone | Yes | 0.9885 | drugbank.ca |
| Acyclovir | Yes |  | PMID: 12878501 |
| Adapalene | Yes | 0.7061 | drugbank.ca |
| Adenosine | Yes | 0.9383 | drugbank.ca |
| Albumin | Yes |  | drugbank.ca |
| Albuterol ipratropium | Yes |  | drugbank.ca |
| Alendronate | Yes | 0.7065 | drugbank.ca |
| Alfuzosin | Yes | 0.6215 | drugbank.ca |
| Allopurinol | Yes | 0.9885 | drugbank.ca |
| Alprazolam | Yes | 0.9794 | drugbank.ca |
| Alteplase | Yes |  | drugbank.ca |
| Aluminum hydroxide | Yes | 0.8181 | drugbank.ca |
| Aluminum magnesium | Yes |  | drugbank.ca |
| Amantadine | Yes | 0.9769 | drugbank.ca |
| Amiloride | Yes | 0.7193 | drugbank.ca |
| Amino acids | Yes |  | PMID: 10736373 |
| Aminocaproic acid | Yes |  | PMID: 9298063 |
| Aminophylline | Yes | 0.5908 | drugbank.ca |
| Amiodarone | Yes | 0.8615 | drugbank.ca |
| Amitriptyline | Yes | 0.9512 | drugbank.ca |
| Ammonium | Yes |  | PMID: 20216550 |
| Ammonium chloride | Yes |  | PMID: 13611039 |
| Ammonium lactate | Yes | 0.7656 | drugbank.ca |
| Amphetamine | Yes | 0.9565 | drugbank.ca |
| Anagrelide | Yes | 0.848 | drugbank.ca |
| Anastrozole | Yes | 0.9382 | drugbank.ca |
| Antipyrine benzocaine | Yes | 0.9931 | drugbank.ca |
| Antipyrine benzocaine glycerin | Yes | 0.9931 | drugbank.ca |
| Aripiprazole | Yes | 0.992 | drugbank.ca |
| Astemizole | Yes | 0.765 | drugbank.ca |
| Atropine | Yes | 0.9569 | drugbank.ca |
| Azelastine | Yes | 0.982 | drugbank.ca |
| Baclofen | Yes | 0.9339 | drugbank.ca |
| Beclomethasone | Yes | 0.9851 | drugbank.ca |
| Belladonna alkaloids opium | Yes |  | https://doi.org/10.1016/B978-0-12-415813-9.00014-3 |
| Belladonna alkaloids phenobarbital | Yes |  | https://doi.org/10.1016/B978-0-12-415813-9.00014-3 |
| Benzocaine | Yes | 0.9609 | drugbank.ca |
| Benzocaine menthol | Yes |  | drugbank.ca |
| Benzonatate | Yes | 0.9355 | drugbank.ca |
| Benzquinamide hydrochloride | Yes | 0.9383 | drugbank.ca |
| Benztropine | Yes | 0.9929 | drugbank.ca |
| Betamethasone | Yes | 0.9781 | drugbank.ca |
| Bethanechol | Yes | 0.9789 | drugbank.ca |
| Bicarbonate | Yes | 0.936 | drugbank.ca |
| Bisacodyl | Yes |  | drugbank.ca |
| Bismuth subsalicylate | Yes | 0.9387 | drugbank.ca |
| Bretylium | Yes | 0.9484 | drugbank.ca |
| Brimonidine | Yes | 0.717 | drugbank.ca |
| Brinzolamide | Yes | 0.8754 | drugbank.ca |
| Brompheniramine | Yes | 0.9576 | drugbank.ca |
| Brompheniramine dextromethorphan pseudoephedrine | Yes | 0.9576 | drugbank.ca |
| Brompheniramine pseudoephedrine | Yes | 0.9576 | drugbank.ca |
| Budesonide | Yes | 0.9533 | drugbank.ca |
| Bupivacaine | Yes | 0.936 | drugbank.ca |
| Bupivacaine hydrochloride epinephrine bitartrate | Yes | 0.936 | drugbank.ca |
| Bupivacine epinephrine | Yes | 0.936 | drugbank.ca |
| Buprenorphine | Yes | 0.9401 | drugbank.ca |
| Bupropion | Yes | 0.975 | drugbank.ca |
| Buspirone | Yes | 0.9839 | drugbank.ca |
| Butoconazole nitrate | Yes | 0.9716 | drugbank.ca |
| Butorphanol | Yes | 0.9463 | drugbank.ca |
| Butorphanol tartrate | Yes | 0.9463 | drugbank.ca |
| Cabergoline | Yes | 0.9383 | drugbank.ca |
| Calcitriol | Yes | 0.8524 | drugbank.ca |
| Calcium | Yes |  | drugbank.ca |
| Calcium acetate | Yes | 0.9601 | drugbank.ca |
| Calcium chloride | Yes | 0.975 | drugbank.ca |
| Calcium cholecalciferol | Yes | 0.959 | drugbank.ca |
| Calcium gluceptate | Yes | 0.5646 | drugbank.ca |
| Calcium vitamin d | Yes |  | drugbank.ca |
| Captopril | Yes | 0.6467 | drugbank.ca |
| Carbamazepine | Yes | 0.9958 | drugbank.ca |
| Carbamide peroxide | Yes |  | PMID: 10882779 |
| Carbidopa levodopa | Yes |  | PMID: 29295587 |
| Carbonic acid | Yes |  |  |
| Carisoprodol | Yes | 0.9679 | drugbank.ca |
| Celecoxib | Yes | 0.9713 | drugbank.ca |
| Cetirizine | Yes | 0.7576 | drugbank.ca |
| Chloral hydrate | Yes | 0.9555 | drugbank.ca |
| Chloramphenicol | Yes | 0.9366 | drugbank.ca |
| Chlordiazepoxide | Yes | 0.9758 | drugbank.ca |
| Chlorhexidine | Yes | 0.7872 | drugbank.ca |
| Chlorhexidine gluconate | Yes | 0.7872 | drugbank.ca |
| Chlorpheniramine | Yes | 0.962 | drugbank.ca |
| Chlorpheniramine methscopolamine phenylephrine | Yes | 0.962 | drugbank.ca |
| Chlorpromazine | Yes | 0.9795 | drugbank.ca |
| Chlorthalidone | Yes | 0.5447 | drugbank.ca |
| Chlorzoxazone | Yes | 0.991 | drugbank.ca |
| Cholecalciferol | Yes | 0.959 | drugbank.ca |
| Ciclopirox | Yes | 0.9892 | drugbank.ca |
| Cilostazol | Yes | 0.9909 | drugbank.ca |
| Ciprofloxacin | Yes |  | drugbank.ca |
| Cisapride | Yes | 0.9383 | drugbank.ca |
| Cisatracurium | Yes | 0.9291 | drugbank.ca |
| Citalopram | Yes | 0.9729 | drugbank.ca |
| Clomipramine | Yes | 0.9793 | drugbank.ca |
| Clonazepam | Yes | 0.9718 | drugbank.ca |
| Clonidine | Yes | 0.9402 | drugbank.ca |
| Clopidogrel | Yes | 0.9847 | drugbank.ca |
| Clorazepate | Yes | 0.8373 | drugbank.ca |
| Clotrimazole | Yes | 0.9837 | drugbank.ca |
| Cocaine | Yes | 0.8805 | drugbank.ca |
| Codeine | Yes | 0.9979 | drugbank.ca |
| Colchicine | Yes | 0.7865 | drugbank.ca |
| Conivaptan | Yes | 0.9522 | drugbank.ca |
| Conjugated estrogens | Yes | 0.9486 | drugbank.ca |
| Cromolyn sodium | Yes | 0.5988 | drugbank.ca |
| Cyclobenzaprine | Yes | 0.9512 | drugbank.ca |
| Cyclophosphamide | Yes | 0.971 | drugbank.ca |
| Cyproheptadine | Yes | 0.986 | drugbank.ca |
| Dantrolene | Yes | 0.9581 | drugbank.ca |
| Dapsone | Yes | 0.9705 | drugbank.ca |
| Darifenacin | Yes | 0.9989 | drugbank.ca |
| Desflurane | Yes | 0.9941 | drugbank.ca |
| Desipramine | Yes | 0.9854 | drugbank.ca |
| Desonide | Yes | 0.9382 | drugbank.ca |
| Desvenlafaxine | Yes | 0.7722 | drugbank.ca |
| Dexamethasone | Yes | 0.9781 | drugbank.ca |
| Dexmedetomidine | Yes | 0.9663 | drugbank.ca |
| Dextroamphetamine | Yes | 0.9565 | drugbank.ca |
| Dextromethorphan | Yes | 1.9565 | drugbank.ca |
| Dextromethorphan guaifenesin | Yes | 2.9565 | drugbank.ca |
| Dextromethorphan guaifenesin pseudoephedrine | Yes | 3.9565 | drugbank.ca |
| Dextrose | Yes | 0.5569 | drugbank.ca |
| Dezocine | Yes | 0.9403 | drugbank.ca |
| Diazepam | Yes | 0.9934 | drugbank.ca |
| Dibucaine | Yes | 0.9876 | drugbank.ca |
| Diclofenac | Yes | 0.9541 | drugbank.ca |
| Dicyclomine | Yes | 0.9769 | drugbank.ca |
| Diltiazem | Yes | 0.506 | drugbank.ca |
| Dimenhydrinate | Yes | 0.8551 | drugbank.ca |
| Diphenhydramine | Yes | 0.9381 | drugbank.ca |
| Docosanol | Yes | 0.9579 | drugbank.ca |
| Dolasetron | Yes | 0.9403 | drugbank.ca |
| Donepezil | Yes | 0.9953 | drugbank.ca |
| Dorzolamide | Yes | 0.6697 | drugbank.ca |
| Doxacurium chloride | Yes | 0.9419 | drugbank.ca |
| Doxazosin | Yes | 0.9627 | drugbank.ca |
| Doxepin | Yes | 0.9381 | drugbank.ca |
| Doxycycline | Yes | 0.9881 | drugbank.ca |
| Dronabidol | Yes | 0.9685 | drugbank.ca |
| Droperidol | Yes | 0.9602 | drugbank.ca |
| Drospirenone | Yes | 0.9383 | drugbank.ca |
| Duloxetine | Yes | 0.9804 | drugbank.ca |
| Dutasteride | Yes | 0.9884 | drugbank.ca |
| Econazole | Yes | 0.9823 | drugbank.ca |
| Entecavir | Yes | 0.873 | drugbank.ca |
| Ephedrine | Yes | 0.5638 | drugbank.ca |
| Epoetin | Yes |  | drugbank.ca |
| Ergocalciferol | Yes | 0.9428 | drugbank.ca |
| Escitalopram | Yes | 0.9729 | drugbank.ca |
| Estazolam | Yes | 0.9796 | drugbank.ca |
| Estradiol | Yes | 0.8917 | drugbank.ca |
| Estrogen | Yes | 0.8917 | drugbank.ca |
| Eszopiclone | Yes | 0.9382 | drugbank.ca |
| Ethanol | Yes | 0.9539 | drugbank.ca |
| Etidronate | Yes | 0.8901 | drugbank.ca |
| Etodolac | Yes | 0.9065 | drugbank.ca |
| Etomidate | Yes | 0.95 | drugbank.ca |
| Ezetimibe | Yes | 0.9074 | drugbank.ca |
| Ezetimibe simvastatin | Yes | 0.9074 | drugbank.ca |
| Famiciclovir | Yes | 0.9429 | drugbank.ca |
| Famotidine | Yes | 0.9382 | drugbank.ca |
| Fenofibrate | Yes | 0.9334 | drugbank.ca |
| Fentanyl | Yes | 0.9901 | drugbank.ca |
| Filgrastim | Yes |  | drugbank.ca |
| Finasteride | Yes | 0.9777 | drugbank.ca |
| Fluconazole | Yes | 0.9382 | drugbank.ca |
| Fludrocortisone | Yes | 0.9731 | drugbank.ca |
| Flumazenil | Yes | 0.9382 | drugbank.ca |
| Fluocinonide | Yes | 0.981 | drugbank.ca |
| Fluorometholone | Yes | 0.9716 | drugbank.ca |
| Fluoxetine | Yes | 0.983 | drugbank.ca |
| Fluphenazine | Yes | 0.975 | drugbank.ca |
| Flurazepam | Yes | 0.9898 | drugbank.ca |
| Folic acid | Yes | 0.7609 | drugbank.ca |
| Fosphenytoin | Yes | 0.9215 | drugbank.ca |
| Fusidic acid | Yes | 0.7909 | drugbank.ca |
| Gabapentin | Yes | 0.9382 | drugbank.ca |
| Glimepiride | Yes | 0.7322 | drugbank.ca |
| Glipizide | Yes | 0.5599 | drugbank.ca |
| Glutamine | Yes | 0.9604 | drugbank.ca |
| Glycopyrrolate | Yes | 0.8436 | drugbank.ca |
| Glycopyrronium | Yes | 0.8436 | drugbank.ca |
| Griseofulvin | Yes | 0.9208 | drugbank.ca |
| Haloperidol | Yes | 0.9465 | drugbank.ca |
| Heparin | Yes |  | drugbank.ca |
| Hydralazine | Yes | 0.9487 | drugbank.ca |
| Hydrochlorothiazide | Yes | 0.9659 | drugbank.ca |
| Hydrocodone | Yes | 0.9984 | drugbank.ca |
| Hydrocortisone | Yes | 0.9383 | drugbank.ca |
| Hydromorphone | Yes | 0.9931 | drugbank.ca |
| Hydroxyurea | Yes | 0.9382 | drugbank.ca |
| Hydroxyzine | Yes | 0.9516 | drugbank.ca |
| Ibuprofen | Yes | 0.9619 | drugbank.ca |
| Imipenem | Yes | 0.9711 | drugbank.ca |
| Imipramine | Yes | 0.9865 | drugbank.ca |
| Imiquimod | Yes | 0.955 | drugbank.ca |
| Indapamide | Yes | 0.8868 | drugbank.ca |
| Indomethacin | Yes | 0.9381 | drugbank.ca |
| Ipratropium | Yes | 0.8883 | drugbank.ca |
| Ipratropium bromide | Yes | 0.8883 | drugbank.ca |
| Irbesartan | Yes | 0.9271 | drugbank.ca |
| Iron | Yes | 0.9733 | drugbank.ca |
| Isoflurane | Yes | 0.994 | drugbank.ca |
| Isoniazid | Yes | 0.9895 | drugbank.ca |
| Isopropyl alcohol | Yes | 0.9694 | drugbank.ca |
| Isoproterenol | Yes | 0.9705 | drugbank.ca |
| Isosorbide dinitrate | Yes | 0.9559 | drugbank.ca |
| Isosorbide mononitrate | Yes | 0.9355 | drugbank.ca |
| Ketamine | Yes | 0.9826 | drugbank.ca |
| Ketoconazole | Yes | 0.6704 | drugbank.ca |
| Ketoprofen | Yes | 0.9382 | drugbank.ca |
| Ketorolac | Yes | 0.7918 | drugbank.ca |
| Ketotifen | Yes | 0.9923 | drugbank.ca |
| Lacosamide | Yes | 0.7181 | drugbank.ca |
| Lactulose | Yes | 0.6609 | drugbank.ca |
| Lamotrigine | Yes | 0.9382 | drugbank.ca |
| Lansoprazole | Yes | 0.7007 | drugbank.ca |
| Levetiracetam | Yes | 0.9821 | drugbank.ca |
| Levocarnitine | Yes | 0.8568 | drugbank.ca |
| Levomepromazine | Yes | 0.9935 | drugbank.ca |
| Levorphanol | Yes | 0.9953 | drugbank.ca |
| Levothyroxine | Yes | 0.6886 | drugbank.ca |
| Lidocaine | Yes |  | drugbank.ca |
| Lidocaine epinephrine | Yes |  | drugbank.ca |
| Lidocaine prilocaine | Yes |  | drugbank.ca |
| Lindane | Yes | 0.9862 | drugbank.ca |
| Linezolid | Yes | 0.9363 | drugbank.ca |
| Lithium | Yes | 0.9708 | drugbank.ca |
| Loperamide | Yes | 0.7709 | drugbank.ca |
| Loratadine | Yes | 0.9754 | drugbank.ca |
| Loratadine pseudoephedrine | Yes |  | drugbank.ca |
| Lorazepam | Yes | 0.9641 | drugbank.ca |
| Lovastatin | Yes | 0.9287 | drugbank.ca |
| Lubiprostone | Yes | 0.8984 | drugbank.ca |
| Magnesium citrate | Yes |  | PMID: 29920013 |
| Magnesium oxide | Yes | 0.9837 | drugbank.ca |
| Magnesium sulfate | Yes | 0.9646 | drugbank.ca |
| Mecamylamine | Yes | 0.9771 | drugbank.ca |
| Meclizine | Yes | 0.9656 | drugbank.ca |
| Medroxyprogesterone acetate | Yes | 0.9617 | drugbank.ca |
| Megestrol acetate | Yes | 0.9617 | drugbank.ca |
| Melatonin | Yes | 0.9928 | drugbank.ca |
| Menthol | Yes | 0.9408 | drugbank.ca |
| Meperidine | Yes | 0.9876 | drugbank.ca |
| Mepivacaine | Yes | 0.9749 | drugbank.ca |
| Metacycline | Yes | 0.9833 | drugbank.ca |
| Metaxalone | Yes | 0.9747 | drugbank.ca |
| Metformin | Yes | 0.5868 | drugbank.ca |
| Methadone | Yes | 0.9772 | drugbank.ca |
| Methocarbamol | Yes | 0.5747 | drugbank.ca |
| Methylnaltrexone | Yes | 0.9268 | drugbank.ca |
| Methylphenidate | Yes | 0.9663 | drugbank.ca |
| Methylprednisolone | Yes | 0.9484 | drugbank.ca |
| Metoclopramide | Yes | 0.9713 | drugbank.ca |
| Metocurine iodide | Yes | 0.9374 | drugbank.ca |
| Metolazone | Yes | 0.5944 | drugbank.ca |
| Metronidazole | Yes | 0.9297 | drugbank.ca |
| Mexiletine | Yes | 0.9382 | drugbank.ca |
| Miconazole | Yes | 0.9823 | drugbank.ca |
| Midazolam | Yes | 0.9724 | drugbank.ca |
| Milnacipran | Yes | 0.9889 | drugbank.ca |
| Minoxidil | Yes | 0.9496 | drugbank.ca |
| Mirtazapine | Yes | 0.9855 | drugbank.ca |
| Misoprostol | Yes | 0.938 | drugbank.ca |
| Mivacurium chloride | Yes | 0.9465 | drugbank.ca |
| Modafinil | Yes | 0.9947 | drugbank.ca |
| Mometasone | Yes | 0.9721 | drugbank.ca |
| Montelukast | Yes | 0.9311 | drugbank.ca |
| Morphine | Yes | 0.9882 | drugbank.ca |
| Morphine sulfate | Yes | 0.9882 | drugbank.ca |
| Mycophenolate | Yes | 0.5826 | drugbank.ca |
| Nabumetone | Yes |  | drugbank.ca |
| Nalbuphine | Yes | 0.9642 | drugbank.ca |
| Naloxone | Yes | 0.9787 | drugbank.ca |
| Naphazoline | Yes | 0.9693 | drugbank.ca |
| Naproxen | Yes | 0.6881 | drugbank.ca |
| Nefazodone | Yes | 0.9744 | drugbank.ca |
| Neostigmine | Yes | 0.9583 | drugbank.ca |
| Niacin | Yes | 0.9648 | drugbank.ca |
| Nicotine | Yes | 0.9748 | drugbank.ca |
| Nitazoxanide | Yes | 0.7175 | drugbank.ca |
| Nitrofurantoin | Yes | 0.958 | drugbank.ca |
| Nitroglycerin | Yes | 0.9113 | drugbank.ca |
| Nitroprusside | Yes | 0.9314 | drugbank.ca |
| Nitrous oxide | Yes | 0.9872 | drugbank.ca |
| Nizatidine | Yes | 0.5 | drugbank.ca |
| Norepinephrine | Yes | 0.9762 | drugbank.ca |
| Nortriptyline | Yes | 0.9478 | drugbank.ca |
| Nystatin | Yes | 0.9581 | drugbank.ca |
| Nystatin triamcinolone | Yes |  | drugbank.ca |
| Olanzapine | Yes | 0.9652 | drugbank.ca |
| Olopatadine | Yes | 0.6925 | drugbank.ca |
| Ondansetron | Yes | 0.9882 | drugbank.ca |
| Oxandrolone | Yes | 0.9537 | drugbank.ca |
| Oxaprozin | Yes | 0.9512 | drugbank.ca |
| Oxazepam | Yes | 0.9641 | drugbank.ca |
| Oxcarbazepine | Yes | 0.9975 | drugbank.ca |
| Oxybutynin | Yes | 0.9418 | drugbank.ca |
| Oxycodone | Yes | 0.9885 | drugbank.ca |
| Oxymorphone | Yes | 0.9382 | drugbank.ca |
| Pamidronate | Yes | 0.6175 | drugbank.ca |
| Pancuronium bromide | Yes | 0.878 | drugbank.ca |
| Pantoprazole | Yes | 0.795 | drugbank.ca |
| Paricalcitol | Yes | 0.795 | drugbank.ca |
| Paroxetine | Yes | 0.9869 | drugbank.ca |
| Pemoline | Yes | 0.983 | drugbank.ca |
| Penciclovir | Yes | 0.9386 | drugbank.ca |
| Pentobarbital | Yes | 0.9713 | drugbank.ca |
| Permethrin | Yes | 0.8736 | drugbank.ca |
| Phenazopyridine | Yes | 0.874 | drugbank.ca |
| Phenobarbital | Yes | 0.9736 | drugbank.ca |
| Phenol | Yes | 0.9076 | drugbank.ca |
| Phenoxybenzamine | Yes | 0.9629 | drugbank.ca |
| Phenylpropanolamine | Yes | 0.5843 | drugbank.ca |
| Phenytoin | Yes | 0.976 | drugbank.ca |
| Physostigmine | Yes | 0.9975 | drugbank.ca |
| Phytonadione | Yes | 0.8941 | drugbank.ca |
| Pioglitazone | Yes | 0.8753 | drugbank.ca |
| Pipecuronium | Yes | 0.6351 | drugbank.ca |
| Pirbuterol | Yes | 0.9442 | drugbank.ca |
| Polymyxin b | Yes | Drubank | drugbank.ca |
| Potassium chloride | Yes | 0.975 | drugbank.ca |
| Potassium iodide | Yes | 0.9702 | drugbank.ca |
| Prazosin | Yes | 0.9479 | drugbank.ca |
| Prednisolone | Yes | 0.9383 | drugbank.ca |
| Prednisolone acetate | Yes |  | drugbank.ca |
| Prednisone | Yes | 0.9383 | drugbank.ca |
| Pregabalin | Yes | 0.9382 | drugbank.ca |
| Prilocaine | Yes | 0.9285 | drugbank.ca |
| Probenecid | Yes | 0.5486 | drugbank.ca |
| Prochlorperazine | Yes | 0.9781 | drugbank.ca |
| Prochlorperazine maleate | Yes |  | drugbank.ca |
| Progesterone | Yes | 0.982 | drugbank.ca |
| Promazine | Yes | 0.987 | drugbank.ca |
| Promethazine | Yes | 0.9855 | drugbank.ca |
| Propafenone | Yes | 0.958 | drugbank.ca |
| Propantheline | Yes | 0.9012 | drugbank.ca |
| Propofol | Yes | 0.9381 | drugbank.ca |
| Propoxyphene | Yes | 0.9503 | drugbank.ca |
| Propylene glycol | Yes | 0.7184 | drugbank.ca |
| Pseudoephedrine | Yes | 0.5638 | drugbank.ca |
| Pyridostigamine | Yes | 0.9818 | drugbank.ca |
| Pyridoxine | Yes | 0.6889 | drugbank.ca |
| Quazepam | Yes | 0.9789 | drugbank.ca |
| Quetiapine | Yes | 0.9906 | drugbank.ca |
| Quinine sulfate | Yes | 0.9382 | drugbank.ca |
| Rabeprazole | Yes | 0.6469 | drugbank.ca |
| Ramelteon | Yes | 0.9934 | drugbank.ca |
| Remifentanil | Yes | 0.9381 | drugbank.ca |
| Risedronate | Yes | 0.9172 | drugbank.ca |
| Risperidone | Yes | 0.9925 | drugbank.ca |
| Rizatriptan | Yes | 0.9406 | drugbank.ca |
| Rocuronium | Yes | 0.8569 | drugbank.ca |
| Ropinirole | Yes | 0.9971 | drugbank.ca |
| Salicylic acid | Yes | 0.6165 | drugbank.ca |
| Salmeterol | Yes | 0.8862 | drugbank.ca |
| Salsalate | Yes | 0.8946 | drugbank.ca |
| Scopolamine | Yes | 0.8218 | drugbank.ca |
| Selegiline | Yes | 0.9809 | drugbank.ca |
| Selenium sulfide | Yes | 0.9786 | drugbank.ca |
| Sertraline | Yes | 0.9713 | drugbank.ca |
| Sevelamer | Yes | 0.9723 | drugbank.ca |
| Sevoflurane | Yes | 0.9941 | drugbank.ca |
| Sildenafil | Yes | 0.6461 | drugbank.ca |
| Silver sulfadiazine | Yes | 0.9438 | drugbank.ca |
| Simvastatin | Yes | 0.9422 | drugbank.ca |
| Sitagliptin | Yes | 0.9438 | drugbank.ca |
| Sodium bicarbonate | Yes | 0.936 | drugbank.ca |
| Solifenacin | Yes | 0.6159 | drugbank.ca |
| Spironolactone | Yes | 0.932 | drugbank.ca |
| Succinylcholine | Yes | 0.7717 | drugbank.ca |
| Sucralfate | Yes | 0.8803 | drugbank.ca |
| Sufentanil | Yes | 0.9886 | drugbank.ca |
| Sulfacetamide | Yes | 0.9881 | drugbank.ca |
| Sulfacetamide sodium urea | Yes | 0.9881 | drugbank.ca |
| Sulfamethoxazole | Yes | 0.9382 | drugbank.ca |
| Sulindac | Yes | 0.8325 | drugbank.ca |
| Sumatriptan | Yes | 0.9626 | drugbank.ca |
| Tacrolimus | Yes | 0.9659 | drugbank.ca |
| Tadalafil | Yes | 0.7821 | drugbank.ca |
| Tamoxifen | Yes | 0.5838 | drugbank.ca |
| Tamsulosin | Yes | 0.5915 | drugbank.ca |
| Telmisartan | Yes | 0.8794 | drugbank.ca |
| Temazepam | Yes | 0.9745 | drugbank.ca |
| Terazosin | Yes | 0.9646 | drugbank.ca |
| Terbinafine | Yes | 0.9381 | drugbank.ca |
| Terconazole | Yes | 0.6657 | drugbank.ca |
| Terfenadine | Yes | 0.5319 | drugbank.ca |
| Testosterone | Yes | 0.973 | drugbank.ca |
| Theophylline | Yes | 0.9902 | drugbank.ca |
| Thiamine | Yes | 0.933 | drugbank.ca |
| Thiopental | Yes | 0.9505 | drugbank.ca |
| Thioridazine | Yes | 0.9901 | drugbank.ca |
| Thrombin | Yes |  | drugbank.ca |
| Tiagabine | Yes | 0.9382 | drugbank.ca |
| Tiotropium | Yes | 0.5208 | drugbank.ca |
| Tizanidine | Yes | 0.8592 | drugbank.ca |
| Tolbutamide | Yes | 0.9321 | drugbank.ca |
| Tolnaftate | Yes | 0.9689 | drugbank.ca |
| Tolterodine | Yes | 0.9194 | drugbank.ca |
| Topiramate | Yes | 0.9382 | drugbank.ca |
| Torasemide | Yes | 0.6871 | drugbank.ca |
| Tramadol | Yes | 0.9382 | drugbank.ca |
| Trazodone | Yes | 0.9829 | drugbank.ca |
| Triamcinolone | Yes | 0.9528 | drugbank.ca |
| Triamterene hydrochlorothiazide | Yes | 0.8735 | drugbank.ca |
| Triazolam | Yes | 0.9719 | drugbank.ca |
| Trifluoperazine | Yes | 0.9814 | drugbank.ca |
| Trimethobenzamide | Yes | 0.8121 | drugbank.ca |
| Trimethoprim | Yes | 0.9381 | drugbank.ca |
| Tropicamide | Yes | 0.9777 | drugbank.ca |
| Ubiquinone | Yes |  | PMID: 27383171 |
| Ursodiol | Yes | 0.9288 | drugbank.ca |
| Valacyclovir | Yes | 0.9389 | drugbank.ca |
| Valganciclovir | Yes | 0.9383 | drugbank.ca |
| Valproic acid | Yes | 0.9626 | drugbank.ca |
| Varenicline | Yes | 0.9908 | drugbank.ca |
| Vecuronium bromide | Yes | 0.878 | drugbank.ca |
| Venlafaxine | Yes | 0.9382 | drugbank.ca |
| Verapamil | Yes | 0.6323 | drugbank.ca |
| Vitamin a | Yes | 0.9665 | drugbank.ca |
| Vitamin a d | Yes | 0.9665 | drugbank.ca |
| Vitamin b | Yes |  | drugbank.ca |
| Vitamin b c | Yes | 0.8532 | drugbank.ca |
| Vitamin b1 | Yes | 0.933 | drugbank.ca |
| Vitamin c | Yes | 0.8532 | drugbank.ca |
| Vitamin d | Yes |  | PMID: 21872806 |
| Vitamin e | Yes | 0.9767 | drugbank.ca |
| Vitamin k | Yes | 0.8941 | drugbank.ca |
| Vitamin k3 | Yes | 0.8265 | drugbank.ca |
| Voriconazole | Yes | 0.9047 | drugbank.ca |
| Warfarin | Yes | 0.9124 | drugbank.ca |
| Xylometazoline hcl | Yes | 0.7293 | drugbank.ca |
| Zaleplon | Yes | 0.9751 | drugbank.ca |
| Zinc | Yes | 0.9733 | drugbank.ca |
| Zinc oxide | Yes |  | drugbank.ca |
| Zinc sulfate monohydrate | Yes |  | drugbank.ca |
| Ziprasidone | Yes | 0.9759 | drugbank.ca |
| Zolpidem | Yes | 0.9392 | drugbank.ca |
| Zonisamide | Yes | 0.9755 | drugbank.ca |
