## Supplementary material for "Pharmacological Management of Acute Spinal Cord Injury: A longitudinal multi-cohort observational study": Table e7

| **Supplementary Table 7: List of drugs that were indicated prophylactical or preventative reasons** | | |
| --- | --- | --- |
| **Drug** | **Organ system** | **Details** |
| **Indication was labelled as 'prophylaxis'** | | |
| acetaminophen | General disorders and administration site conditions | Fever and headache prophylaxis |
| acetaminophen | General disorders and administration site conditions | Fever prophylaxis |
| acetaminophen | Pain | Pain prophylaxis |
| acetaminophen | Gastrointestinal disorders | Ulcer prophylaxis |
| acetylcysteine | Respiratory, thoracic and mediastinal disorders | Respiratory prophylaxis |
| albuterol | Respiratory, thoracic and mediastinal disorders | Pulmonary prophylaxis |
| albuterol | Respiratory, thoracic and mediastinal disorders | Respiratory prophylaxis |
| albuterol | Respiratory, thoracic and mediastinal disorders | Wheezing prophylaxis |
| alprazolam | Psychiatric disorders | Anxiety prophylaxis |
| aluminum magnesium | Gastrointestinal disorders | Ulcer prophylaxis |
| aluminum magnesium simethicone | Gastrointestinal disorders | Ulcer prophylaxis |
| aminophylline | Respiratory, thoracic and mediastinal disorders | Pulmonary prophylaxis |
| amoxicillin | Infections and infestations | Infection prophylaxis |
| amoxicillin clavulanate potassium | Infections and infestations | Infection prophylaxis |
| ampicillin | Infections and infestations | Infection prophylaxis |
| ampicillin sulbactam | Infections and infestations | Infection prophylaxis |
| acetylsalicylic acid | Vascular disorders | Deep vein thrombosis prophylaxis |
| atropine | Cardiac disorders | Bradycardia prophylaxis |
| aztreonam | Infections and infestations | Infection prophylaxis |
| bacitracin | Infections and infestations | Infection prophylaxis |
| benztropine | Nervous system disorders | Prophylaxis |
| capsaicin | Gastrointestinal disorders | Ulcer prophylaxis |
| carbamazepine | Nervous system disorders | Anxiety prophylaxis |
| cefadroxil | Infections and infestations | Uti prophylaxis |
| cefamandole | Infections and infestations | Infection prophylaxis |
| cefazolin | Infections and infestations | Infection prophylaxis |
| cefoperazone | General disorders and administration site conditions | Prophylaxis after temp |
| cefotaxime | Infections and infestations | Infection prophylaxis |
| cefotetan | Infections and infestations | Prophylaxis |
| cefoxitin | Infections and infestations | Infection prophylaxis |
| ceftazidime | Infections and infestations | Infection prophylaxis |
| ceftizoxime | Surgical and medical procedures | Post-op infection prophylaxis |
| cephapirin | Infections and infestations | Prophylaxis |
| chloral hydrate | Surgical and medical procedures | Prophylaxis |
| chlordiazepoxide | Vascular disorders | Deep vein thrombosis prophylaxis |
| chlorpromazine | Psychiatric disorders | Seizure prophylaxis |
| choline magnesium trisalicylate | General disorders and administration site conditions | Prophylaxis |
| cimetidine | Gastrointestinal disorders | Gastric reflux prophylaxis |
| cimetidine | Gastrointestinal disorders | Ulcer prophylaxis |
| ciprofloxacin | Infections and infestations | Infection prophylaxis |
| ciprofloxacin | Infections and infestations | Pneumonia prophylaxis |
| ciprofloxacin | Infections and infestations | Uti prophylaxis |
| cisapride | Gastrointestinal disorders | Esophageal reflux prophylaxis |
| cisapride | Gastrointestinal disorders | Gastric reflux prophylaxis |
| cisapride | Gastrointestinal disorders | Ulcer prophylaxis |
| clindamycin | Respiratory, thoracic and mediastinal disorders | Aspiration prophylaxis |
| clindamycin | General disorders and administration site conditions | Fever prophylaxis |
| clindamycin | Infections and infestations | Infection prophylaxis |
| clindamycin | Infections and infestations | Respiratory prophylaxis |
| cloxacillin | Infections and infestations | Infection prophylaxis |
| cloxacillin | Surgical and medical procedures | Post surgical prophylaxis |
| cloxacillin | Infections and infestations | Post-op infection prophylaxis |
| cloxacillin | Infections and infestations | Pre-op infection prophylaxis |
| dexamethasone | Infections and infestations | Anti-inflammatory prophylaxis |
| dexamethasone | General disorders and administration site conditions | Edema prophylaxis |
| dexamethasone | Surgical and medical procedures | Extubation prophylaxis |
| diazepam | Psychiatric disorders | Confusion, agitation proph. |
| diphenhydramine | Immune system disorders | Allergic reaction prophylaxis |
| diphenhydramine | Immune system disorders | Antihistamine prophylaxis |
| diphenhydramine | Blood and lymphatic system disorders | Blood transfer reaction prophylaxis |
| diphenhydramine | Blood and lymphatic system disorders | Infection prophylaxis |
| diphenhydramine | Skin and subcutaneous tissue disorders | Itching prophylaxis |
| diphenhydramine | Surgical and medical procedures | Pre-op infection prophylaxis |
| diptheria, pertussis, tetanus vaccine | Infections and infestations | Diptheria, pertussis, and tetanus PROPHYLAXIS |
| docusate senna | Gastrointestinal disorders | Constepation prophylaxis |
| enoxaparin | Vascular disorders | Deep vein thrombosis prophylaxis |
| enoxaparin | Vascular disorders | Vascular prophylaxis |
| ephedrine | Vascular disorders | Orthostatic hypothesion prophylaxis |
| epinephrine sulfate | Respiratory, thoracic and mediastinal disorders | Prophylaxis |
| erythromycin | Infections and infestations | Infection prophylaxis |
| etidronate | Musculoskeletal and connective tissue disorders | Ossification heterotopic prophylaxis |
| famotidine | Gastrointestinal disorders | Gastric reflux prophylaxis |
| famotidine | Gastrointestinal disorders | Ulcer prophylaxis |
| fentanyl | Surgical and medical procedures | Pe prophylaxis |
| ferrous sulfate | Blood and lymphatic system disorders | Prophylactic |
| fluconazole | Infections and infestations | Fungal infection prophylaxis |
| fluconazole | Infections and infestations | Yeast infection prophylaxis |
| folic acid | Metabolism and nutrition disorders | Prophylaxis |
| furosemide | Cardiac disorders | Congestive heart failure prophylaxis |
| gentamicin | Infections and infestations | Infection prophylaxis |
| gentamicin | Surgical and medical procedures | Or-related infections prophylaxis |
| gentamicin | Infections and infestations | Sepsis prophylaxis |
| gentamicin | Infections and infestations | Wound infection prophylaxis |
| heparin | Vascular disorders | Deep vein thrombosis prophylaxis |
| heparin | Vascular disorders | Thrombophlebitis prophylaxis |
| heparin | Vascular disorders | Vascular prophylaxis |
| hydrocortisone | Infections and infestations | Infection prophylaxis |
| hydroxyzine | Gastrointestinal disorders | Nausea prophylaxis |
| imipenem cilastatin | Infections and infestations | Uti prophylaxis |
| influenza vaccine | Infections and infestations | Flu prophylaxis |
| iothalamate meglumine | Surgical and medical procedures | Pe prophylaxis |
| ipratropium | Respiratory, thoracic and mediastinal disorders | Pulmonary prophylaxis |
| ketorolac | Pain | Pain prophylaxis |
| lactobacillus | Gastrointestinal disorders | Diarrhea prophylaxis |
| lorazepam | General disorders and administration site conditions | Delirium prophylaxis |
| magnesium hydroxide | Gastrointestinal disorders | Prophylaxis |
| magnesium sulfate | Metabolism and nutrition disorders | Hypomagnesemia prophylaxis |
| methadone | Pain | Prophylactic |
| methocarbamol | Nervous system disorders | Prophylaxis |
| methylprednisolone | Surgical and medical procedures | Post-op prophylaxis |
| metoclopramide | Gastrointestinal disorders | Nausea prophylaxis |
| metoclopramide | Surgical and medical procedures | Pre-op infection prophylaxis |
| metoclopramide | Gastrointestinal disorders | Ulcer prophylaxis |
| metronidazole | Infections and infestations | Infection prophylaxis |
| miconazole | Blood and lymphatic system disorders | Platelet aggregation prophylaxis |
| midazolam | Psychiatric disorders | Anxiety prophylaxis |
| midazolam | Pain | Pain prophylaxis |
| misoprostol | Gastrointestinal disorders | Stress ulcer prophylaxis |
| multivitamins | Metabolism and nutrition disorders | Prophylaxis |
| nafcillin | Infections and infestations | Infection prophylaxis |
| nafcillin | Surgical and medical procedures | Pre-op infection prophylaxis |
| neomycin | Infections and infestations | Prophylaxis |
| nicotine | Psychiatric disorders | Nicotine withdrawal prophylaxis |
| nifedipine | Vascular disorders | Prophylaxis |
| nitrofurantoin | Infections and infestations | Uti prophylaxis |
| nizatidine | Gastrointestinal disorders | Ulcer prophylaxis |
| nystatin | Infections and infestations | Infection prophylaxis |
| ofloxacin | Infections and infestations | Infection prophylaxis |
| omeprazole | Gastrointestinal disorders | Stress ulcer prophylaxis |
| penicillin | Infections and infestations | Infection prophylaxis |
| phenytoin | Nervous system disorders | Seizures prophylaxis |
| phytonadione | Blood and lymphatic system disorders | Prophylaxis |
| piperacillin | Infections and infestations | Prophylaxis |
| pneumococcal vaccine | Infections and infestations | Prophylactic |
| potassium chloride | Metabolism and nutrition disorders | Potassium prophylaxis |
| povidone iodine | Infections and infestations | Infection prophylaxis |
| prednisone | Infections and infestations | Infection prophylaxis |
| prochlorperazine | Gastrointestinal disorders | Nausea prophylaxis |
| promethazine | Gastrointestinal disorders | Nausea prophylaxis |
| promethazine | Pain | Pain prophylaxis |
| pseudoephedrine | Psychiatric disorders | Dizziness prophylaxis |
| ranitidine | Gastrointestinal disorders | Ulcer prophylaxis |
| red blood cells | Blood and lymphatic system disorders | Low hematocrit prophylaxis |
| sertraline | Psychiatric disorders | Prophylaxis |
| simethicone | Gastrointestinal disorders | Ulcer prophylaxis |
| sodium bicarbonate | Metabolism and nutrition disorders | Low bicarbonate prophylaxis |
| sucralfate | Gastrointestinal disorders | Ulcer prophylaxis |
| sulfamethoxazole | Infections and infestations | Uti prophylaxis |
| tetanus vaccine | Infections and infestations | Tetanus prophylaxis |
| tetracycline | Infections and infestations | Prophylaxis |
| thrombin | Blood and lymphatic system disorders | Bleeding prophylaxis |
| ticarcillin | Infections and infestations | Prophylaxis |
| tolterodine | Renal and urinary system disorders | Prophylaxis |
| trimethoprim | Infections and infestations | Uti prophylaxis |
| vancomycin | Infections and infestations | Infection prophylaxis |
| vitamin B1 | Metabolism and nutrition disorders | Prophylaxis |
| vitamin K | Blood and lymphatic system disorders | Prophylaxis |
| warfarin | Vascular disorders | Deep vein thrombosis prophylaxis |
| warfarin | Vascular disorders | Vascular prophylaxis |
| **Indication was labelled as 'prevention'** | | |
| acetaminophen | General disorders and administration site conditions | Prevent fever |
| acetaminophen hydrocodone | Pain | Prevent pain |
| acetylcysteine | Respiratory, thoracic and mediastinal disorders | Prevent congestion |
| acyclovir | Infections and infestations | Prevent herpes outbreak |
| albuterol | Respiratory, thoracic and mediastinal disorders | Prevent bronchospasm |
| albuterol | Respiratory, thoracic and mediastinal disorders | Prevent congestion |
| aluminum magnesium | Gastrointestinal disorders | Prevent ulcers |
| aluminum magnesium simethicone | Pain | Prevent pain |
| amitriptyline | Psychiatric disorders | Prevent depression |
| ampicillin sulbactam | Infections and infestations | Prevent infections |
| bacitracin | Infections and infestations | Prevent infections |
| bisacodyl | Gastrointestinal disorders | Prevent constipation |
| calcium gluconate | Metabolism and nutrition disorders | Prevent electrolyte imbalance |
| carbamazepine | Nervous system disorders | Prevent seizures |
| cefazolin | General disorders and administration site conditions | Prevent infections |
| cefotaxime | Infections and infestations | Prevent infections |
| ceftazidime | Infections and infestations | Prevent infections |
| cephalexin | Infections and infestations | Prevent infections |
| chlordiazepoxide | Vascular disorders | Deep venous thrombosis |
| chlorpromazine | Psychiatric disorders | Prevent shivering |
| cimetidine | Gastrointestinal disorders | Prevent ulcers |
| ciprofloxacin | Infections and infestations | Prevent infections |
| cisapride | Gastrointestinal disorders | Prevent constipation |
| cisapride | Gastrointestinal disorders | Prevent heartburn |
| cisapride | Gastrointestinal disorders | Prevent ulcers |
| clotrimazole | Infections and infestations | Prevent yeast infection |
| diazepam | Psychiatric disorders | Prevent anxiety |
| diphenhydramine | Blood and lymphatic system disorders | Prevent reaction from vit. K |
| docusate | Gastrointestinal disorders | Prevent constipation |
| enoxaparin | Vascular disorders | Deep venous thrombosis |
| ephedrine | Vascular disorders | Prevent neurogenic hypotension |
| famotidine | Gastrointestinal disorders | Prevent ulcers |
| iron | Blood and lymphatic system disorders | Prevent anemia |
| fludrocortisone | Nervous system disorders | Prevent swelling |
| folic acid | Blood and lymphatic system disorders | Prevent deep vein thrombosis |
| furosemide | Vascular disorders | Prevent fld overload |
| gentamicin | Infections and infestations | Prevent infections |
| heparin | Vascular disorders | Prevent deep vein thrombosis |
| hydroxyzine | Gastrointestinal disorders | Prevent nausea |
| influenza vaccine | Infections and infestations | Prevent flu |
| lorazepam | Psychiatric disorders | Prevent anxiety |
| magnesium sulfate | Metabolism and nutrition disorders | Prevent electrolyte imbalance |
| magnesium sulfate | Metabolism and nutrition disorders | Prevent hypomagnesemia |
| magnesium oxide | Metabolism and nutrition disorders | Prevent hypomagnesemia |
| mannitol | Nervous system disorders | Prevent spinal cord swelling |
| meperidine | Pain | Prevent pain |
| metoclopramide | Gastrointestinal disorders | Prevent constipation |
| metoclopramide | Gastrointestinal disorders | Prevent nausea |
| metoclopramide | Gastrointestinal disorders | Prevent ulcers |
| metronidazole | Infections and infestations | Prevent infections |
| miconazole | Infections and infestations | Prevent yeast infection |
| misoprostol | Gastrointestinal disorders | Prevent ulcers |
| morphine | Pain | Prevent Pain |
| multivitamin supplements | Metabolism and nutrition disorders | Prevent deep vein thrombosis |
| nalbuphine | Gastrointestinal disorders | Prevent nausea |
| nicotine | Psychiatric disorders | Prevent nicotine withdrawal |
| nystatin | Infections and infestations | Prevent infections |
| ondansetron | Gastrointestinal disorders | Prevent nausea |
| oxacillin | Infections and infestations | Prevent infections |
| phenobarbital | Nervous system disorders | Prevent seizures |
| phenytoin | Nervous system disorders | Prevent seizures |
| plasma | Blood and lymphatic system disorders | Prevent bleeding |
| potassium chloride | Metabolism and nutrition disorders | Prevent hypokalemia |
| potassium phosphate | Metabolism and nutrition disorders | Prevent electrolyte imbalance |
| potassium phosphate | Metabolism and nutrition disorders | Prevent hypophosphatemia |
| potassium chloride | Metabolism and nutrition disorders | Prevent electrolyte imbalance |
| promethazine | Gastrointestinal disorders | Prevent nausea |
| promethazine | General disorders and administration site conditions | Prevent itching |
| ranitidine | Gastrointestinal disorders | Prevent ulcers |
| sodium phosphate | Metabolism and nutrition disorders | Prevent hypophosphatemia |
| sodium bicarbonate | Metabolism and nutrition disorders | Prevent electrolyte imbalance |
| sodium chloride | Metabolism and nutrition disorders | Prevent electrolyte imbalance |
| sodium phosphate | Metabolism and nutrition disorders | Prevent electrolyte imbalance |
| sucralfate | Gastrointestinal disorders | Prevent ulcers |
| sulfamethoxazole | Infections and infestations | Prevent infections |
| tetanus vaccine | Infections and infestations | Prevent tetanus |
| tolterodine | Infections and infestations | Prevent incontinence |
| trimethobenzamide | Gastrointestinal disorders | Prevent nausea |
| trimethoprim | Infections and infestations | Prevent infections |
| vitamin K | Blood and lymphatic system disorders | Prevent bleeding |
| vitamin B1 | Metabolism and nutrition disorders | Prevent deep vein thrombosis |
| warfarin | Vascular disorders | Prevent deep vein thrombosis |
| zinc oxide | Skin and subcutaneous tissue disorders | Prevent skin breakdown |
